# Brain-Organ Hypersynchrony and Cognitive Decline in Alzheimer’s Disease: Potential Links with Tauopathy and Glymphatic Dysfunction

**DOI:** 10.64898/2026.04.22.26351474

**Authors:** Luoyu Wang, Lianghua Li, Yitian Tao, Yifan Jia, Juan Yue, Yajuan Zhang, Yihan Wang, Yan Zhang, Mei Xin, Jianjun Liu, Feng Shi, Chenpeng Zhang, Han Zhang

## Abstract

Alzheimer’s disease (AD) is increasingly recognized to have systemic physiological correlates alongside central neurodegeneration. Here, we explored brain-organ network (BON) connectivity in AD (n=28) and healthy controls (n=23) using time-resolved quasi-dynamic analysis of plateau-phase total-body ^18^F-tau-PET. We found that AD-related pathophysiology was linked not only to cerebral tau aggregation, but also to altered signal synchronization across the brain-organ network, despite comparable body tracer distribution. Network topology analyses revealed the occipitotemporal cortex and the spinal cord as key nodes in this altered systemic network. Furthermore, exploratory mediation analyses demonstrated that BON dysregulation is cross-sectionally linked to cognitive deficits, with statistical associations observed for both cortical tau burden and imaging markers of impaired glymphatic clearance. This total-body PET study provides first-ever direct evidence repositioning AD as a multi-organ disorganization disease. These findings provide a novel framework for investigating brain-body interactions and systemic vulnerabilities in neurodegenerative disorders.

## Introduction

Alzheimer’s disease (AD) is a progressive neurodegenerative disorder characterized by extracellular β-amyloid (Aβ) plaques and intraneuronal neurofibrillary tangles composed of hyperphosphorylated tau in the brain, which are associated with synaptic dysfunction and irreversible neuronal loss^1,2^. While Aβ deposition is typically regarded as an early initiating event, pathological tau accumulation correlates more strongly with clinical symptom severity and disease progression^3,4^. Despite contemporary therapeutic and strategies targeting these core brain pathologies, disease-modifying therapies have yielded limited clinical benefit^5,6^. The long-standing therapeutic challenges underscore a critical limitation in current understanding and suggest paralleled pathogenetic mechanisms extending beyond the brain. Consequently, the traditional view of AD as a brain-centric disorder may overlook emerging evidence of its broader, systemic involvement of body organs^7–9^.

Recent research has increasingly challenged the long-held brain-centric view of AD by demonstrating that it could also involve extracranial organs via immune^7^, metabolic^10^, and lymphatic pathways^9,11^. These studies have identified several brain-organ interactions, such as brain-heart^9^, -gut^8^or -liver^12^ axes, while others have also linked peripheral conditions including cardiovascular diseases, microbial metabolites, and hepatic dysfunction to cognitive decline^12,13^. In addition, glymphatic malfunction has become another hot topic of the brain-body involvement in AD, as the glymphatic system modulates cerebrospinal-interstitial exchange, refreshing the brain environment and facilitating the communication between the brain and extracranial organs^14^. Collectively, this evidence leads to an increasingly important hypothesis that cross-body, systemic pathological changes could be an integral, yet historically underexplored component of AD pathogenesis. Despite the progress, a key limitation that makes this hypothesis far from elucidating is that all the previous studies only conducted anatomically isolated investigations, without directly capturing the discoordination across multiple organ systems in AD. A holistic, network-level view of brain-organ interactions become urgently necessary for AD study^15^.

The above limitation is primarily due to the drawbacks of traditional imaging techniques: they cannot capture dynamic information of the entire body due to the static nature of acquisition and the difficulty of synchronized cross-organ imaging coverage. Recent pioneering human brain-organ interactions studies inevitably fail to fully account for extensive physiological interconnections among all major organ systems in the whole body^15^. Total-body PET/CT is a novel imaging tool that provides technical feasibility of capturing dynamic, multi-organ biophysiology concurrently, making the investigation of aberrant systemic interactions in AD possible, as it uniquely enables simultaneous quantification of tracer distribution across the entire body in a single imaging session^16^. While most PET studies provide only a static “snapshot” at tracer equilibrium^17^, the exceptional sensitivity and signal-to-noise ratio of the total-body PET allows for the reconstruction of time-resolved, dynamic “frames” ^18,19^. While early-phase total-body PET^18,19^, captures rapid tracer delivery and specific binding kinetics, time-resolved reconstructions of the late (plateau) phase offer a unique opportunity to detect low-frequency systemic physiological and hemodynamic fluctuations. Conceptually analogous to resting-state functional MRI (rs-fMRI), the temporal covariance of these late-phase physiological signals across distant body regions can be used to map macroscopic physiological synchronization, constructing a systemic brain-organ network (BON).

Building on the evidence of systemic involvement in AD pathogenesis and the emerging ability of measuring systemic physiological synchronization using total-body PET, we hypothesize that the disease manifests not only as central tau proteinopathy, but also through altered systemic physiological coordination, reflected in an aberrant BON. This network dysregulation may signify an erosion of systemic autonomic or hemodynamic resilience that normally supports neuroprotective mechanisms. To test this, we developed a novel human-based BON quantification framework using dynamic total-body tau-PET for comprehensive brain-organ interaction mapping based on the correlated late-phase tracer kinetics derived from major brain regions and extracranial organs. By comparing the BON between AD and healthy controls, we characterized altered patterns of systemic physiological synchronization in AD. Furthermore, using exploratory mediation analyses, we identified cross-sectional statistical associations suggesting that BON dysfunction is linked to cognitive decline, exacerbated tau pathology, and imaging markers of glymphatic impairment. Rather than viewing AD strictly as an isolated brain disease, our findings highlight complex systemic physiological alterations accompanying the disease, revealing the BON as a promising frontier for investigating holistic disease markers and systemic vulnerabilities in neurodegeneration.

## Results

### Study cohort

Our data were derived from a single-center, retrospective cohort with Aβ-, tau-, and FDG-PET, and multimodal brain MRI using PET/MR and total-body PET/CT, comprising 28 patients with AD and 23 age-, sex-, and education-matched negative controls (NC). The diagnosis of AD was established according to the 2024 NIA-AA criteria, requiring positivity for both amyloid-β (Aβ) and tau PET, supported by characteristic hypometabolism on ^18^F-FDG-PET. The NC group was negative for both Aβ and tau pathologies. The multimodal brain MRI included 3D T1-weighted and T2-FLAIR sequences, and, where available, other advanced MRI sequences such as rs-fMRI and diffusion tensor imaging (DTI) (**Supplementary Fig. 1**). Demographic and clinical characteristics are summarized in **Table 1**. See detailed inclusion/exclusion criteria and imaging protocols in **Methods**.

**Table 1.**
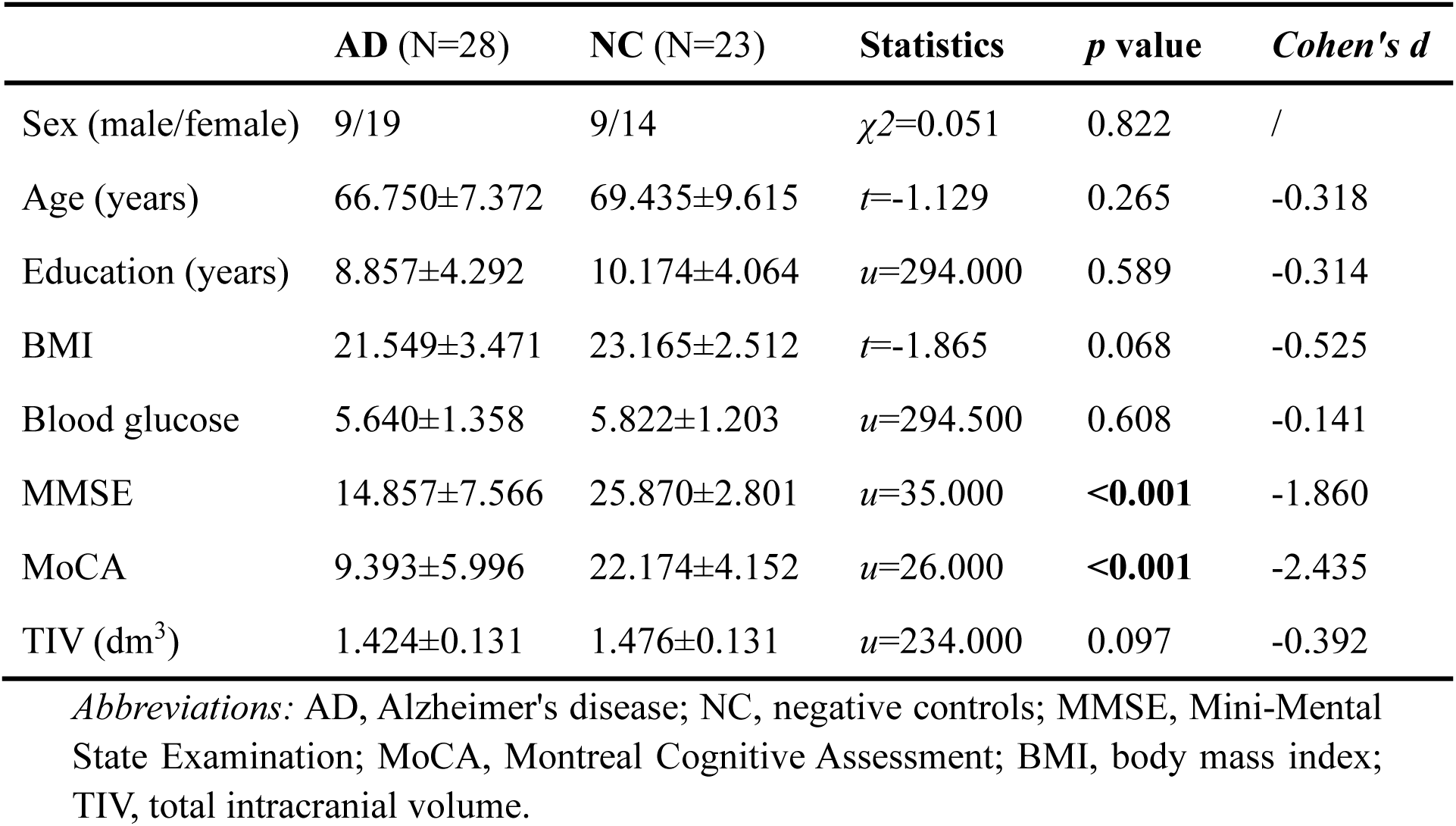
Demographic and clinical characteristics of the AD and NC groups.

### Elevated brain-organ connectivity in AD

To characterize spatiotemporal synchronization of late-phase PET signals, which likely reflect systemic physiological and hemodynamic fluctuations, across the entire body, we established a BON construction and comparison framework, which integrates dynamic total-body PET/CT reconstruction and high-resolution brain structural MRI, with dedicated motion correction, cross-modality registration, key brain area/body-organ segmentation, standardized uptake value ratio (SUVR) calculation, and BON calculation (detailed flowchart in **Supplementary Fig. 2**). Specifically, averaged dynamic late-phase tracer kinetics time series were extracted from ten bilateral cortical regions (bilateral frontal, temporal, parietal, occipital and insula regions) and nine extracranial regions (major organs and tissues such as kidneys, liver, lungs, colon, spleen, skeleton, heart, spinal cord, and skeletal muscles). After regressing out motion-related signals, BON matrices were generated with Pearson correlation of the regional averaged time series.

Group-level comparisons using the network-based statistic (NBS), controlled for age, sex, and education, revealed that the AD group exhibited significantly stronger brain-organ connectivity compared with the NC group, characterized by a subnetwork of 35 supra-threshold connections (**Fig. 1a**), with nine intracerebral (brain-brain), five inter-organ (organ-organ), and the majority of 21 brain-body connections. We further identified the most affected brain regions and body organs through a subsequent hub analysis. These include the right occipital and temporal cortices, and the spinal cord (**Fig. 1b**). The kidneys, liver, skeleton, and heart were also found as key extracranial regions harboring the identified brain/extracranial hubs (**Fig. 1c**). Network analysis further delineated three categories of such AD-related abnormalities: ***1)*** *intracerebrally*, the connections with the strongest group differences converged on the right hemisphere, with dominant connections identified between the bilateral occipital lobes and within the right frontal-temporal network (**Fig. 1d**); ***2)*** *extracranially,* the spinal cord acted as a central anchor of alterations, exhibiting pronounced coupling changes with the lungs, skeletal muscle, and kidneys (**Fig. 1e**); ***3)*** *brain-extracranial connectivity,* which primarily involved the alterations from the right occipital lobe to the liver, and from right temporal lobe to skeletal muscle and heart, alongside the spinal cord’s altered coupling with the left parietal lobe (**Fig. 1f**).

**Fig. 1.**
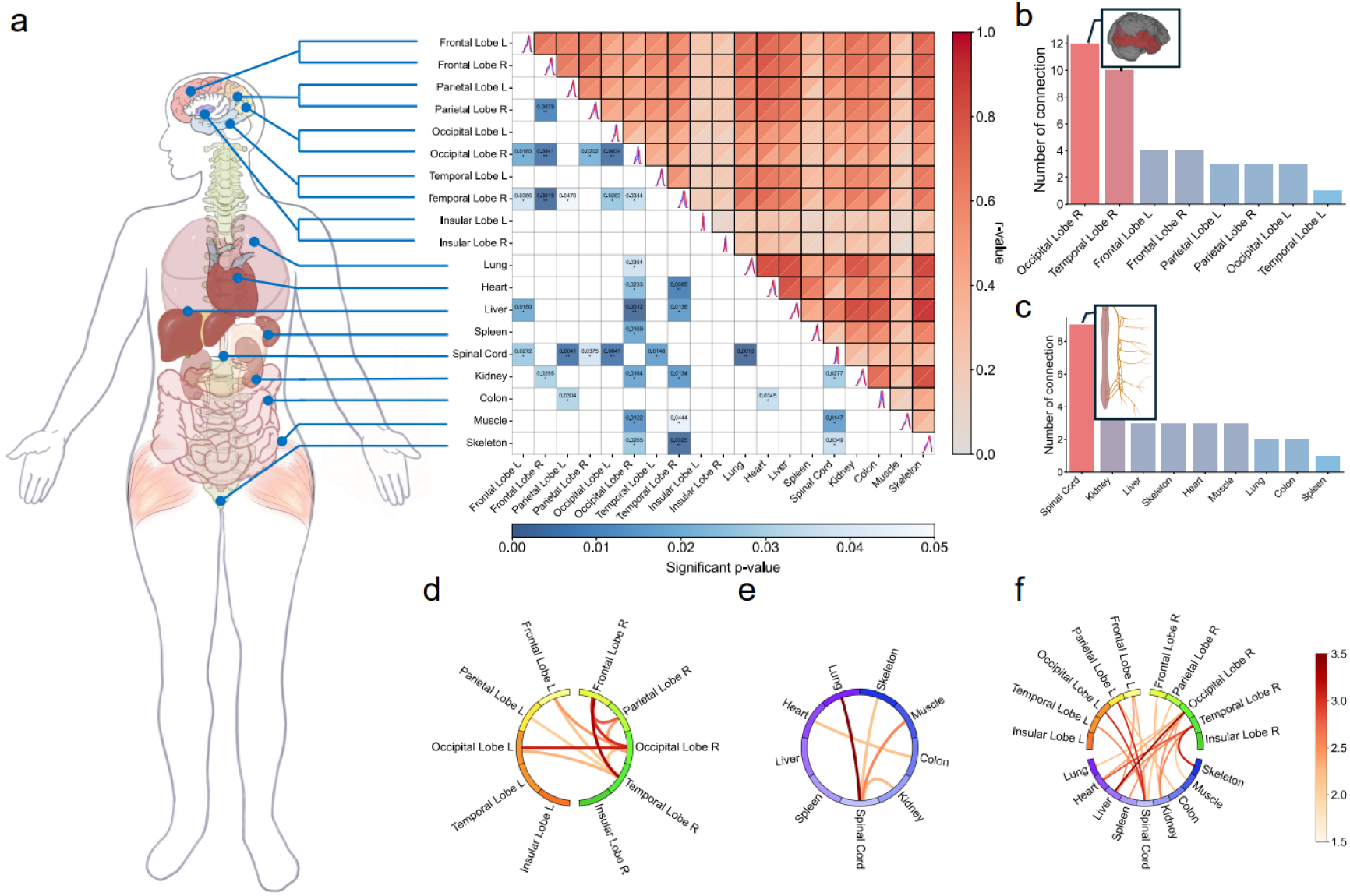
Altered brain-organ connectivity in AD. **a** Case-control heatmap of brain-organ connectivity after adjustment for age, sex, and education. Significance was assessed using the NBS correction (primary edge-wise *p* < 0.05, component-wise *p* < 0.05) to control family-wise error. The upper triangular matrix displays the mean values for the two groups (AD in the top-left, NC in the bottom-right); the lower triangular matrix shows corresponding significance levels. Diagonal plots show the distribution of connection strengths for each node (red, AD; blue, NC). **b-c** Numbers of cerebral regions (**b**) and extracranial regions (**c**) involved in NBS-identified significant connections. **d-f** Chord diagram of significantly altered brain-organ connections, which is categorized into (**d**) intracerebral (brain-brain), (**e**) inter-organ (organ-organ), and (**f**) brain-extracranial (brain-organ) connections. Nodes represent cerebral or extracranial regions, and edge color encodes *t* values. Significance is indicated as \**p* < 0.05, \*\**p* < 0.01. *Abbreviations:* NBS, Network-Based Statistic; AD, Alzheimer’s disease; NC, negative controls.

### Statistical mediation models link altered brain-organ connectivity, cerebral tau burden, and cognitive performance

To quantify tau burden across cerebral and extracranial regions, conventional (static) total-body tau-PET analyses were also carried out to measure normalized SUVRs in the cerebral and extracranial regions (detailed in **Methods**, see also **Supplementary Information** and **Supplementary Fig. 3**). Compared with the NCs, ADs exhibited significantly higher tau SUVRs across all major cortical lobes (**Fig. 2a**; Benjamini-Hochberg false discovery rate [FDR] corrected, *q* < 0.05). However, no significant group differences were observed in tau SUVRs at any of the extracranial regions (**Fig. 2b**). Tau burden in the left occipital, parietal, and temporal cortices was negatively correlated with Mini-Mental State Examination (MMSE) scores, while the left occipital tau burden showed a negative correlation with Montreal Cognitive Assessment (MoCA) scores (**Fig. 2c**; partial Spearman correlation, controlled for age, sex, and education, FDR corrected, *q* < 0.05). The conventional static PET analysis validated cerebral tau accumulation in AD, and its association with cognitive decline, but did not reveal extracranial involvement as dynamic PET did.

**Fig. 2.**
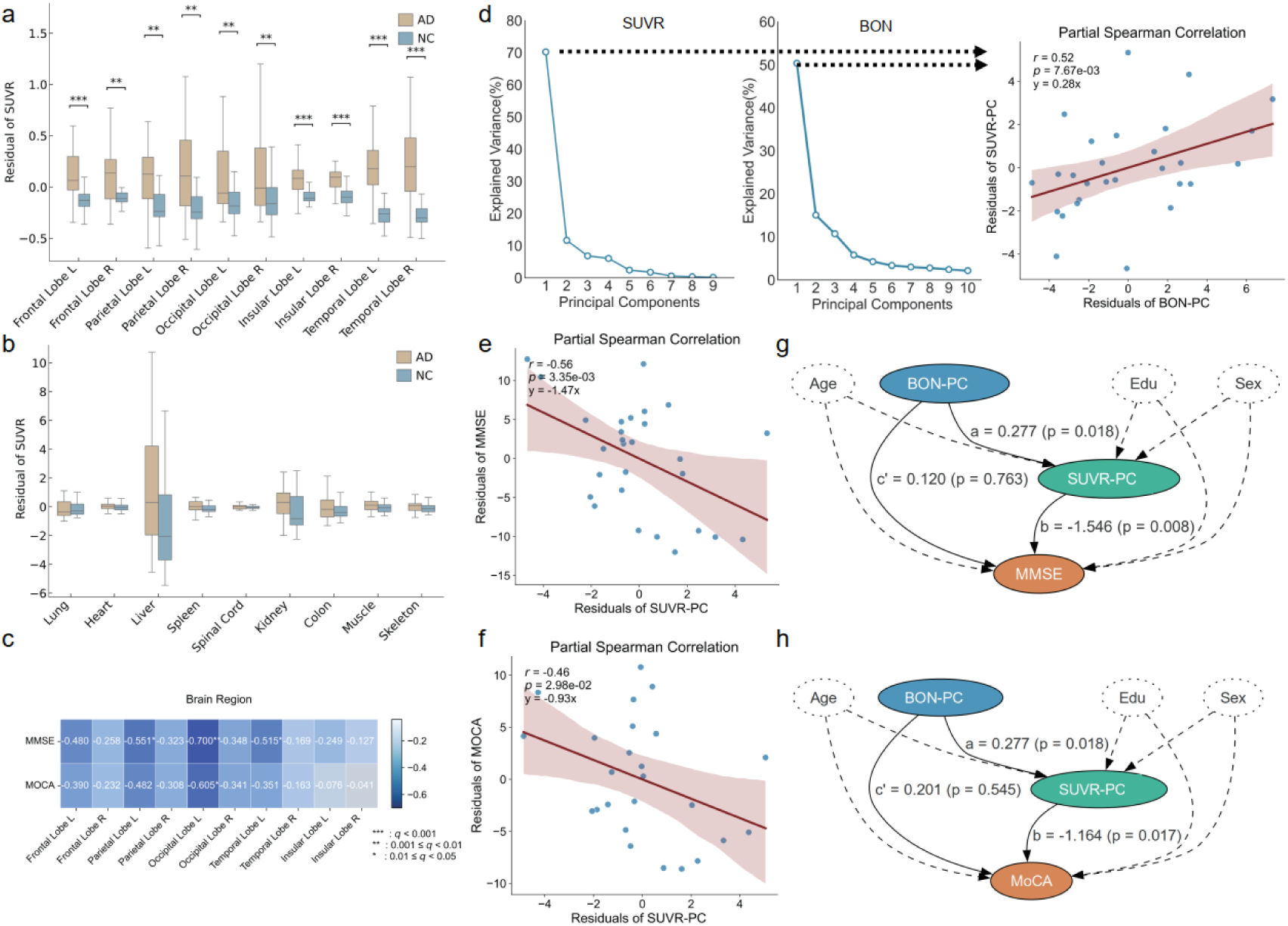
Increased cortical tau burden and its association with brain-organ connectivity and impaired cognitive function. **a–b** Group comparisons of tau tracer uptake (SUVRs) across major cortical lobes (**a**) and extracranial regions **(b**) between the AD and NC groups. SUVRs were normalized to the cerebellar cortex for brain regions and to the aorta for extracranial regions. Group differences were evaluated using general linear models controlling for age, sex, and education, with multiple-comparison correction by Benjamini–Hochberg FDR (two-sided, *q* < 0.05). **c** Partial Spearman correlations between cortical SUVRs and cognitive performance (MMSE and MoCA) within the AD group, controlling for age, sex, and education (FDR-corrected, *q* < 0.05). **d** Probabilistic principal component analysis (PPCA) summarizing distributed tau burden (SUVR-PC) and brain-organ connectivity (BON-PC) components, with their explained variance and partial correlation (*r* = 0.52, *p* = 0.008). **e, f** Partial Spearman correlations showing negative associations of SUVR-PC with MMSE (**e**) and MoCA (**f**). **g, h** Mediation models testing whether cortical tau burden mediates the link between brain-organ connectivity and cognitive function. BON-PC was positively associated with SUVR-PC (*a* = 0.28, *p* = 0.018), which was inversely associated with MMSE (**g,** *b* = −1.55, *p* = 0.008) and MoCA (**h,** *b* = −1.164, *p* = 0.017). Significance is indicated as \**q* < 0.05, \*\**q* < 0.01, \*\*\**q* < 0.001. *Abbreviations:* AD, Alzheimer’s disease; NC, negative controls; FDR, false discovery rate; MMSE, Mini-Mental State Examination; MoCA, Montreal Cognitive Assessment.

Next, we investigated the relationship between the aforementioned static tau burden and the dynamic PET-based BON connectivity strength. To better measure such a high-dimensional correlation, we applied probabilistic principal component analysis (PPCA) separately to the static SUVRs and the BON connections with group differences. This resulted in two composite scores: the first principal component of tau burden (SUVR-PC) and that of brain-organ connectivity (BON-PC), explaining 70% and 50% of the respective variances, respectively. In the AD group, the SUVR-PC and BON-PC showed a moderate positive partial correlation after adjusting for age, sex, and education (*r* = 0.52, *p* = 0.0076; **Fig. 2d**). We also found that elevated SUVR-PC was significantly associated with poorer cognitive performance across the AD patients, as reflected by negative Spearman’s correlations with MMSE (*r* = −0.56, *p* = 0.0035; **Fig. 2e**) and MoCA scores (*r* = −0.46, *p* = 0.0298; **Fig. 2f**), after adjusting for the same demographic variables. Individual correlation values between each BON connection and regional SUVRs were also provided (**Supplementary Fig. 4**).

Given correlations between the static and dynamic measures, and between static or dynamic measures and cognitive status, we further investigated whether cerebral tau burden serves as a mediating factor between altered BON connectivity and cognitive decline. The mediation analysis with structural equation modeling revealed that the BON-PC was positively associated with the SUVR-PC (*a* = 0.277, *p* = 0.018, 95% CI [0.070, 0.470]), and a higher tau burden predicted lower scores on both MMSE (*b* = - 1.546, *p* = 0.008, 95% CI [-3.689, −0.280]; **Fig. 2g**) and MoCA (path *b* = −1.164, *p* = 0.017, 95% CI [-3.025, −0.241]; **Fig. 2h**). Meanwhile, the direct effect (i.e., path *c’*) of the BON-PC on the cognitive function after accounting for the mediator was statistically non-significant for both MMSE (*c’* = 0.120, *p* = 0.763, 95% CI [-0.821, 1.036]) and MoCA (*c’* = 0.201, *p* = 0.545, 95% CI [-0.621, 0.968]). Statistically, this pattern is consistent with a model where systemic physiological dysregulation (BON-PC) shares variance with cognitive impairment indirectly through its association with cerebral tau pathology.

To test the directional specificity of such mediation, we also performed reverse mediation analyses by swapping the independent variable and the mediator. In this alternative model, BON-PC was tested as a mediator of the effect from SUVR-PC to the cognitive scores. Although SUVR-PC was associated with BON-PC (*a* = 0.597, *p* = 0.018, 95% CI [0.154, 1.369]), BON-PC failed to significantly predict cognitive outcomes for either MMSE (*b* = 0.120, *p* = 0.763, 95% CI [-0.820, 1.036]) or MoCA (*b* = 0.201, *p* = 0.545, 95% CI [-0.621, 0.968]). Moreover, the direct effect of SUVR-PC on the cognitive scores remained significant after including BON-PC as a potential mediator (path *c’*; MMSE: *c’* = −1.546, *p* = 0.008, 95% CI [-3.688, −0.280]; MoCA: *c’* = −1.164, *p* = 0.017, 95% CI [-3.024, −0.241]). As such, no significant reverse mediation effects were observed, suggesting that within this specific cross-sectional statistical framework, BON alterations are positioned as an upstream statistical correlate.

### Statistical mediation of the association between brain-organ connectivity and cognitive decline by glymphatic MRI indices

As AD has also been hypothesized to involve impaired glymphatic clearance, we investigated whether altered brain-organ connectivity was associated with impaired glymphatic clearance. To comprehensively measure glymphatic function, we derived 11 metrics (summarized in **Fig. 3a–e**) from multimodal MRI for the same AD patients, including choroid plexus volume fraction (CPVF), perivascular space volume fractions in the whole brain, white matter and basal ganglia (PVSVF, PVSVF-WM, PVSVF-BG), free-water indices (FW-All, FW-WM, FW-BG), analysis-along-perivascular-space indices (lALPS, rALPS, mALPS, representing the left, right, and mean values, respectively), and global blood-oxygen-level-dependent–cerebrospinal fluid coupling (gBOLD-CSF), as detailed in **Supplementary Materials**. PPCA was again performed on the indices to yield composite scores (i.e., the first two principal components, GLY-PC1, GLY-PC2), respectively dominated by the FW-WM and gBOLD-CSF (**Fig. 3g–h**). The same partial correlation analyses revealed that BON-PC was positively associated with both GLY-PC1 (*r* = 0.40, *p* = 0.045; **Fig. 3i**) and GLY-PC2 (*r* = 0.61, *p* = 0.001; **Fig. 3j**).

**Fig. 3.**
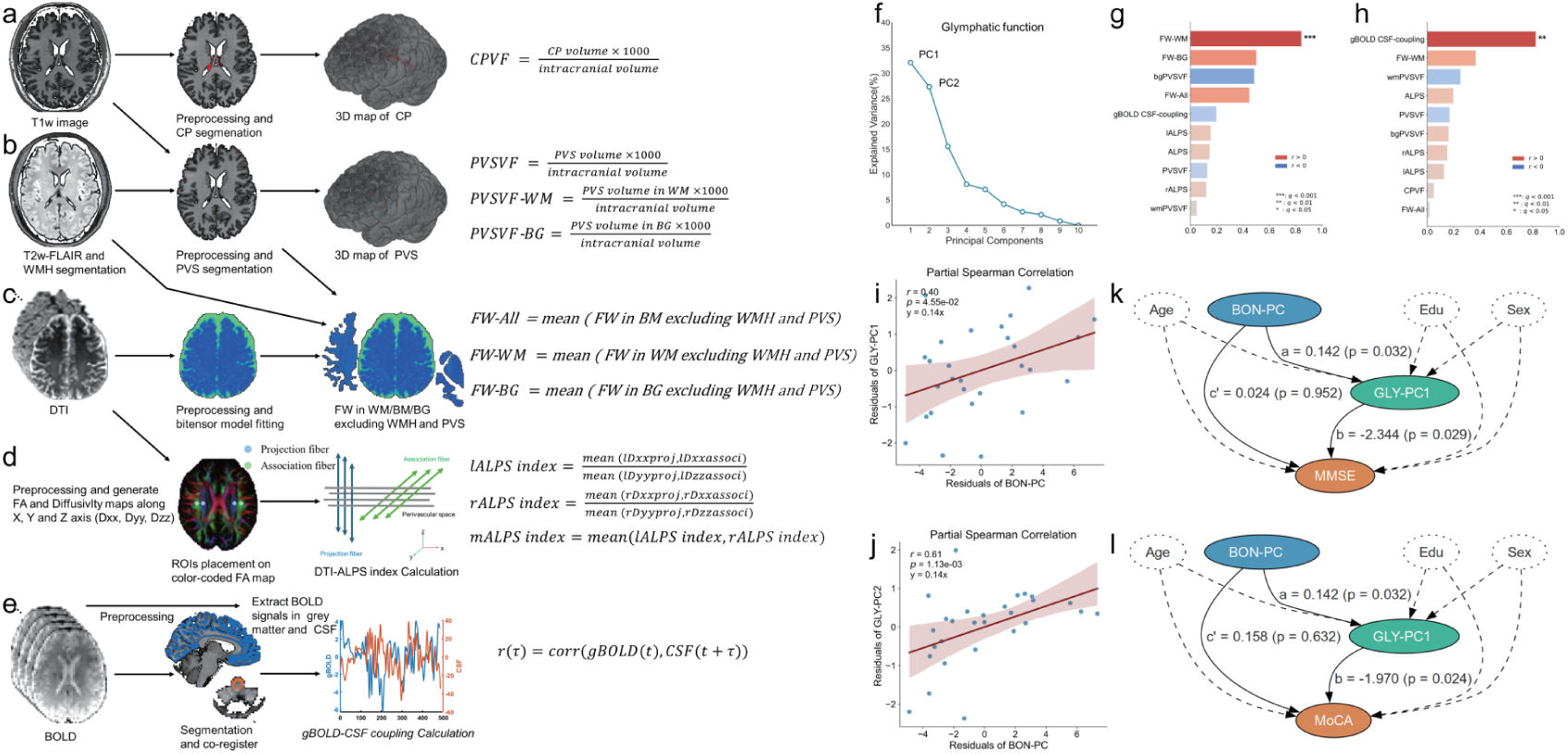
Multimodal MRI-derived glymphatic indices and their association with brain-organ connectivity in AD. **a–e** Workflow for calculating comprehensive glymphatic indices. (**a**) Choroid plexus volume fraction (CPVF) calculated from T1-weighted images. (**b**) Perivascular space volume fractions (PVSVF) computed from T2w/FLAIR, including whole-brain (PVSVF), white-matter (PVSVF-WM), and basal-ganglia (PVSVF-BG) components. (**c**) Free-water indices (FW-All, FW-WM, FW-BG) derived from diffusion tensor imaging. (**d**) Analysis-along-perivascular-space indices (lALPS, rALPS, mALPS, representing the left, right, and mean values, respectively) computed from diffusion tensor imaging (DTI) data. (**e**) Global BOLD-CSF coupling (gBOLD–CSF) estimated from resting-state functional MRI (rs-fMRI) signals. **f** PPCA on the 11 glymphatic indices. GLY-PC1 accounted for 32% of total variance; GLY-PC2 explained 28%. **g–h** Partial Spearman correlations between individual glymphatic indices and (**g**) GLY-PC1 or (**h**) GLY-PC2, adjusted for age, sex, and education. Bars denote correlation magnitude and direction; asterisks indicate significance after Benjamini-Hochberg FDR correction (two-sided, *q* < 0.05). **i–j** Partial Spearman correlations between BON-PC and GLY-PC1/-PC2 in AD adjusted for age, sex and education: (**i**) *r* = 0.40, *p* = 0.045 for BON-PC vs GLY-PC1; (**j**) *r* = 0.61, *p* = 0.001 for BON-PC vs GLY-PC2. Lines indicate least-squares fits; shaded areas represent 95% confidence intervals. **k–l** Mediation models testing whether glymphatic function mediates the link between brain-organ connectivity and cognition. BON-PC was positively associated with GLY-PC1 (*a* = 0.142, *p* = 0.032), which was inversely associated with MMSE (*b* = −2.344, *p* = 0.029) and MoCA (*b* = −1.970, *p* = 0.024). Significance is indicated as \**q* < 0.05, \*\**q* < 0.01, \*\*\**q* < 0.001. *Abbreviations:* AD, Alzheimer’s disease; CPVF, choroid plexus volume fraction; PVSVF, perivascular-space volume fraction; FW, free water; ALPS, analysis along the perivascular space; gBOLD-CSF, global BOLD-CSF coupling; PPCA, probabilistic principal component analysis; NBS, Network-Based Statistic; TIV, total intracranial volume.

To further elucidate the relationship among brain-organ connectivity, glymphatic function, and cognitive function, we performed similar mediation analyses. Consistent with the aforementioned results with tau, glymphatic dysfunction, represented by GLY-PC1, was also found to mediate the relationship between brain-organ connectivity and cognitive scores. Specifically, increased BON-PC correlated with elevated GLY-PC1 (path *a* = 0.142, *p* = 0.032, 95% CI [0.024, 0.290]), which in turn showed an independent cross-sectional association with reduced cognitive scores (MMSE: path *b* = −2.344, *p* = 0.029, 95% CI [-4.481, −0.240], **Fig. 3k**; MoCA: path *b* = −1.970, *p* = 0.024, 95% CI [-3.923, −0.150], **Fig. 3l**). Notably, the direct effect of BON-PC on the cognitive function after accounting for the mediator was non-significant for both MMSE (*c’* = 0.024, *p* = 0.952, 95% CI: [-1.016, 0.788]) and MoCA (*c’* = 0.158, *p* = 0.632, 95% CI: [-0.807, 0.965]). This suggests a significant indirect pathway between BON and cognitive performance through glymphatic function. Exploratory pairwise correlation analyses are detailed in **Supplementary Fig. 5.**

A similar directional-specificity test, treating BON-PC as a mediator of the link from GLY-PC1 to cognitive function, showed that the path from BON-PC to cognitive outcomes was non-significant (MMSE: *b* = 0.024, *p* = 0.952, 95% CI: [-1.016, 0.788]; MoCA: *b* = 0.158, *p* = 0.632, 95% CI: [-0.807, 0.965]), and the direct effects of GLY-PC1 on cognitive function after adding the mediator remained significant (MMSE: *c’* = −2.343, *p* = 0.029, 95% CI: [-4.482, −0.239]; MoCA: *c’* = −1.970, *p* = 0.024, 95% CI: [- 3.922, −0.150]).

### Parallel statistical models associating altered BON, tau burden, glymphatic indices, and cognition

To test whether cerebral tau burden and glymphatic dysfunction are two independent mediators linking abnormally elevated BON to cognitive decline, we also constructed a parallel mediation model, adjusting for age, sex, and education. We found that BON-PC was positively associated with both mediators: SUVR-PC (path *a_1_* = 0.277, *p* = 0.018, 95% CI: [0.070, 0.470]) and GLY-PC1 (path *a_2_* = 0.142, *p* = 0.032, 95% CI: [0.024, 0.290]), which were respectively independently associated with lower MMSE scores (SUVR-PC: path *b_1_* = −1.815, *p* < 0.001, 95% CI: [-3.349, −0.596]; GLY-PC1: path *b_2_* = −2.880, *p* = 0.001, 95% CI: [-4.777, −0.476]). Crucially, the direct effect of BON-PC on MMSE was rendered non-significant (path *c’* = 0.603, *p* = 0.097, 95% CI: [-0.439, 1.270]) after accounting for these mediators, suggesting that the association between BON and MMSE is substantially mediated by these two parallel pathways (**Fig. 4**).

**Fig. 4.**
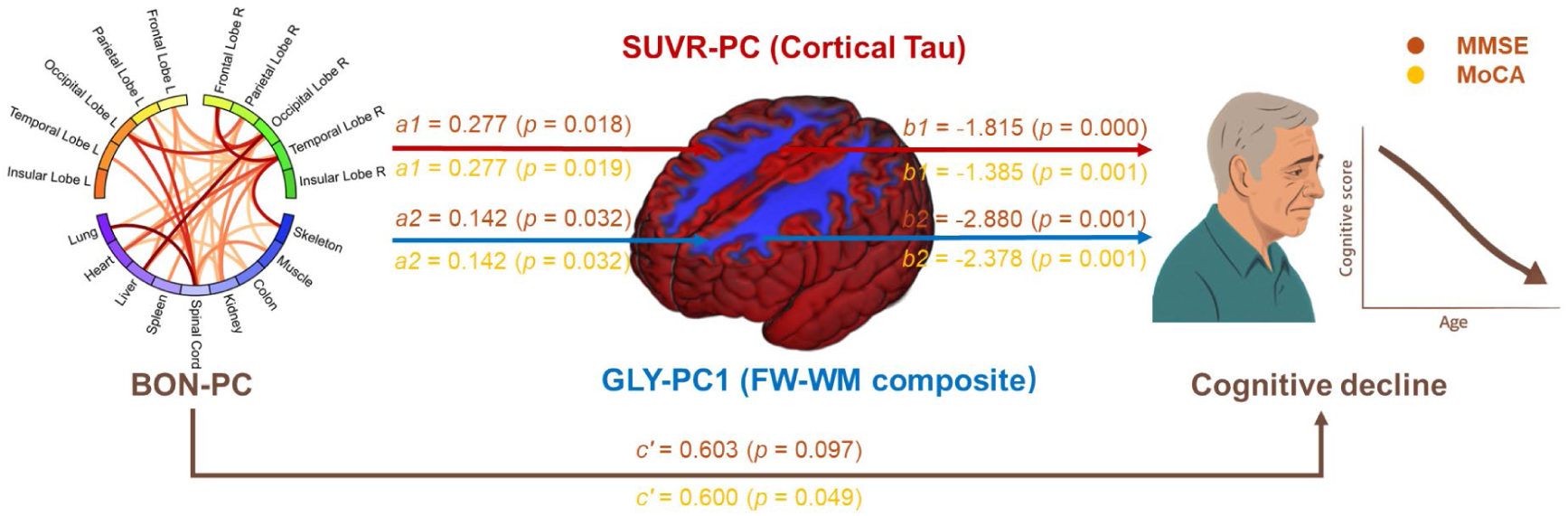
Schematic summary of our proposed parallel mediation model that links brain-organ hyper-synchronization with cognitive decline in AD. The model positions brain-organ connectivity (BON-PC, visualized as a chord diagram on the left) as the predictor of cognitive decline (right) through two independent, non-redundant mechanistic pathways. One pathway is mediated by cortical tau burden (SUVR-PC, red), while the other is mediated by glymphatic dysfunction predominantly in the brain white matter (GLY-PC1, blue). Unstandardized path coefficients and P-values are displayed for each modulatory link (*a_1_*, *a_2_*, *b_1_*, *b_2_*) and for the direct effect (*c’*). The direct effect was non-significant for MMSE and significant for MoCA. The statistics are color-coded to differentiate the two cognitive outcomes: brown text corresponds to the model with MMSE, and yellow text corresponds to the model with MoCA. *Abbreviations:* FW, free water; MMSE, Mini-Mental State Examination; MoCA, Montreal Cognitive Assessment.

Similar patterns were observed for MoCA. BON-PC significantly predicted both SUVR-PC (*a_1_* = 0.277, *p* = 0.019, 95% CI: [0.070, 0.470]) and GLY-PC1 (*a_2_* = 0.142, *p* = 0.032, 95% CI: [0.024, 0.290]). Subsequently, both mediators exhibited significant negative associations with MoCA (SUVR-PC: *b_1_* = −1.385, *p* = 0.001, 95% CI: [-2.759, −0.458]; GLY-PC1: *b_2_* = −2.378, *p* = 0.001, 95% CI: [-4.135, −0.407]). In this model, the direct effect was significant but markedly attenuated after adding the mediators (path *c’* = 0.600, *p* = 0.049, 95% CI: [0.243, 1.287]), indicating a partial mediation effect. Collectively, these findings suggest that systemic physiological alterations (BON hyper-synchronization) share variance with cognitive impairment via two statistically independent profiles: central tau accumulation and imaging markers of glymphatic dysfunction.

Please note that MMSE and MoCA were significantly correlated in this cohort (Pearson’s *r* = 0.853, *p* < 0.001), but they were not identical measures. MMSE is more commonly used to reflect global cognitive impairment and disease severity, whereas MoCA is generally considered more sensitive to milder deficits, particularly in executive and visuospatial domains. Therefore, the observation of comparable mediation patterns across both scales suggests that the association between brain-organ hyper-synchronization and cognition is relatively robust across related but non-identical cognitive measures and may extend across a broader spectrum of cognitive impairment.

## Discussion

AD has been regarded primarily as a neurological disorder and approached using brain-centric imaging. However, the persistent challenges for many brain-targeted therapies indicate that key systemic factors outside the brain are still being largely overlooked. Our study aims to expand the conventional brain-centric view of AD by mapping it as a condition with profound systemic physiological correlates. Leveraging the unprecedented sensitivity and spatiotemporal resolution of total-body tau PET/CT, we provide the first direct characterization of the brain-organ connectivity (BON) based on the inter-regional synchronization of post-plateau, late-phase, low-frequency tracer temporal kinetics, which may reflect systemic hemodynamic oscillations and tracer tissue-exchange dynamics in both cerebral and extracranial tissues. We found that the strength of BON was abnormally elevated in the ADs compared to NCs. From the abnormal BON, we identified right occipitotemporal cortex and spinal cord as dominant hubs orchestrating this pathologically systemic hyper-connectivity. Furthermore, our exploratory statistical models suggest that this BON dysregulation shares variance with cognitive decline via two statistically independent profiles: exacerbated cortical tau burden and imaging markers of impaired glymphatic clearance.

At the systemic level, our findings demonstrate a widespread reorganization of systemic BON coupling that involves both key brain regions and extracranial organs, while tau depositions were still unaltered in the extracranial organs. This finding challenges the conventional neurocentric view of AD pathogenesis using an *in vivo* imaging-based paradigm on human subjects. By analyzing the entire 70-min dynamic total-body tau PET data, distinct temporal SUVR patterns were observed across different tissues (**Supplementary Fig. 7)**. We found that, at approximately 60-70 min post-injection, the SUVR in both cerebral and extracranial compartments stabilized compared to the major kinetic wash-in/wash-out patterns (**Supplementary Fig. 2b**). Physiologically, ultra-slow oscillations in tracer kinetics at the plateau phase may provide valuable information, potentially reflecting vasomotor activity and autonomic rhythms for maintaining a healthy, dynamic, whole-body meta-status flexibility. This could provide a flexible temporal scaffold for inter-organ coordination^20,21^ as measured by the BON. The increased temporal coherence of ultra-slow tau SUVR fluctuations in ADs indicates that such a flexibility might be impaired, manifesting as pathological *hyper-synchronization*. This result might suggest aberrant over-synchronization of vascular and autonomic rhythms associated with tau pathology, because the reduction in the healthy, adaptive systemic variability in ADs well aligns with established AD pathophysiology mechanisms, such as endothelial dysfunction, impaired neurovascular coupling, and autonomic imbalance; all collectively constrain systemic regulatory capacity^22–24^. Another possible explanation is that astrocyte activation (e.g., elevated GFAP) and neuroinflammation in AD likely exacerbate vascular/autonomic dysregulation, reinforcing the pathological synchrony as observed^25,26^. This interpretation remains provisional, but it provides a parsimonious systems-level frame for linking peripheral physiology to central vulnerability and should be further validated using animal models with a prospective design.

Moving to the architecture of this network, we identified the right occipitotemporal cortex and the spinal cord as dominant hubs orchestrating this systemic reorganization, providing a preliminary map of extracranial nodes that might serve as early intervention targets. The right occipitotemporal cortex emerged as a principal disease-related cortical hub. Anatomically, it lies within posterior circulation territories that support slow hemodynamic oscillations and is adjacent to venous and glymphatic-related drainage pathways implicated in interstitial clearance^27,28^. Its right-lateralized prominence may reflect an interaction between asymmetric venous outflow anatomy and AD-related reductions in vascular damping capacity. Given that the right transverse sinus often handles a substantial fraction of venous drainage^29,30^, disease-associated vascular stiffening could plausibly facilitate transmission of systemic low-frequency fluctuations to nearby cortex, contributing to elevated synchrony. Functionally, the occipitotemporal cortex integrates visual, sensory, and memory-related processing and is vulnerable to early tau accumulation, even at the pre-symptomatic stage of AD^31^. The convergence of physical pressure from the venous system and biological vulnerability from tau pathology likely forces this region into a state of pathological synchronization with the systemic environment. In contrast, both sides of the insular cortex that are crucial for autonomic nervous system and cardiovascular regulation^32^ did not exhibit differential connectivity with other brain regions or extracranial organs, suggesting that AD-related hyper-synchronization stems from a bottom-up loss of physiological flexibility rather than a primary failure of central regulatory centers. This decoupling implies that the systemic environment could become too rigid to be effectively buffered by autonomic modulation, even when the regulatory hub itself remains stable.

Within the same reorganized network, the spinal cord emerged as a principal extracranial hub in the altered BON. This likely reflects its role as a conduit for bidirectional communication between the brain, CSF, and systemic circulation. The spinal cord is anatomically positioned to facilitate both downward propagation of pathological tau (consistent with cerebrospinal fluid and blood biomarker findings) and upward transmission of peripheral inflammatory and autonomic feedback signals to the brain^33–35^. In AD, increased coupling between the spinal cord and cortical regions may reflect a heightened transmission of altered systemic autonomic and vascular rhythms to the central nervous system, rather than specific protein spreading. This aligns with the spinal cord’s structural vulnerability to systemic hemodynamic changes in neurodegenerative states.

Beyond these hubs, extracranial organs (including kidney, liver, skeleton, and heart) formed secondary clusters that did not show overt tau deposition changes but nevertheless participated in the altered systemic rhythms. The involvement of these organs is driven by their critical roles in maintaining systemic and cerebral homeostasis. For instance, the liver and kidneys function as essential systemic hubs for Aβ and tau clearance; hepatic impairment is responsible for approximately 60% of Aβ circulation and elimination failures that directly elevates the cerebral amyloid burden^36^. Similarly, renal dysfunction induces systemic inflammation and oxidative stress, which can exacerbate structural damage in the hippocampus and other memory-related regions^37^. Furthermore, cardiovascular manifestations, such as reduced ejection fraction and autonomic imbalance, contribute significantly to AD pathogenesis by altering vascula-hemodynamic signaling and compromising cerebral perfusion^38,39^. Representative cross-system links such as coupling between right occipital regions and liver, or between right temporal regions and skeletal muscle/heart suggest that regions implicated in sensory processing and memory may be embedded within broader physiological coordination patterns rather than operating in isolation. These findings indicate that AD-related pathology can extend to a breakdown in coordinated physiological regulation across the body systems. Taken together, our results of multiple organ involvement well integrates prior findings that reported fragmented organ-axis interactions (e.g. brain-kidneys^40^, brain-liver^12^, brain-bone^41^, brain-heart^9^) and organ-cognition links (e.g. hepatic dysfunction^12,42^ and cardiovascular diseases^13^), more straightforwardly supporting the view of AD as a multi-organ network disorder.

These extracranial organs warrant dedicated future studies for better understanding of AD pathogenesis.

Based on our cross-sectional mediation models linking systemic network alterations to tau accumulation^43^ and glymphatic clearance indices^44^, we propose a hypothesis for future validation: systemic physiological dysregulation may act synergistically with central neurodegeneration. We hypothesize that these coupled associations may reflect a global loss of autonomic flexibility across cerebral and peripheral systems^45,46^. In a healthy state, coordinated autonomic control allows the cerebrovascular system to function as a physiological buffer, shielding the microcirculation from disruptive systemic pressure waves^46^. However, as adaptive control diminishes in AD, hemodynamic forces may begin to drive pathological hypersynchrony across the brain-organ network. On one hand, this unbuffered brain-organ coupling may impose chronic neurovascular stress^47,48^. The direct transmission of systemic fluctuations into vulnerable cortical hubs could create a mechanically and metabolically adverse microenvironment, which might directly facilitate tau aggregation and propagation^49^. On the other hand, the loss of hemodynamic shielding might simultaneously contribute to fluid clearance failure. When systemic pressure transients propagate intracranially, they could compromise the delicate vascular compliance required for efficient paravascular pumping^50,51^.

In addition, our multivariate analysis separated this glymphatic impairment into two distinct axes: interstitial stagnation (GLY-PC1, reflecting free-water retention) and hemodynamic-CSF decoupling (GLY-PC2) ^52,53^. Although heightened brain-organ connectivity correlated with deficits in both domains, our mediation modeling indicated that only interstitial stagnation (GLY-PC1) significantly mediated the pathway to cognitive decline. This specific finding suggests that the physical retention of interstitial fluid and the potentially associated accumulation of metabolic waste might act as the direct driving force of neurodegeneration in this axis. These findings suggest that glymphatic impairment may not merely be a secondary effect of protein aggregation. By concurrently promoting interstitial stagnation and exacerbating tau toxicity, this dual-mechanism framework might illustrate how systemic dysregulation could be directly translated into neural compromise.

Beyond these biological insights, we introduce a scalable, cost-effective methodology that repurposes routine clinical PET scans for high spatiotemporal resolution dynamics and network analysis. By exploiting subtle time-activity curve (TAC) fluctuations during the tracer plateau, we implemented a quasi-dynamic reconstruction data using list-mode derived from standard clinical acquisition protocols^54^. Connection-level reliability of the BON, validated by split-half reproducibility analyses on the dynamic PET, demonstrated robust stability with approximately 85% edge-wise ICC values exceeding 0.4 (**Supplementary Fig. 6)**. This approach constructs comprehensive networks while avoiding the prohibitive costs of full dynamic protocols^55^, transforming widely available clinical data into research-grade insights and opening an implementable pathway for broad clinical adoption.

While providing unprecedented systemic tau mapping, our study has several limitations. The single center study limited its cohort size; future multi-center study should be conducted to ensure population generalizability. The retrospective, cross-sectional design cannot establish causality among BON dysregulation, glymphatic failure, tau spreading, and cognitive decline, which necessitates longitudinal studies on small animals. Glymphatic proxies (e.g., ALPS index, PVS burden) rather than direct CSF measurements might introduce confounding by vascular comorbidities. Off-target tau-PET binding (choroid plexus, skeletal) was mitigated but not eliminated by reference normalization.

In conclusion, all the results culminate in a novel whole-body pathologically maladaptive hyper-synchronization model as promising hypothesis for AD pathogenesis. This model suggests that AD may involve impaired dynamic coupling between the brain and key extracranial regions, and that its loss of flexibility may indicate a dysregulation of the homeostatic control that maintains healthy systemic rigidity, a breakdown that is eventually transduced into central neurodegeneration. We thus advocate an imaging-based, data-supported hypothetical framework for AD: the collapse of regulatory buffers, such as autonomic resilience, might allow unbuffered systemic fluctuations to propagate into the brain, creating an adverse physiological environment that progresses in tandem with tau-protein aggregation and impaired clearance mechanisms. Such cascading processes may provide valuable interventional insights in the future. Crucially, our study also introduces a scalable methodology to repurpose routine clinical PET data for research into high spatiotemporal resolution dynamics and networks by re-analyzing late-phase kinetics from standard clinical time windows using quasi-dynamic reconstruction. This methodology offers a cost-effective avenue and opens up a new direction for the recent hot research topic of total-body PET studies, enabling its implementation in broader clinical settings.

## Methods

### Participants and clinical information

Cohort composition, imaging acquisitions, and collected clinical information are summarized in **Supplementary Fig. 1**. This single-site, retrospective study included 51 participants recruited at Renji Hospital (Shanghai, China) between June 2021 and January 2024, comprising 28 AD and 23 age-, sex-, and education-matched NCs. The diagnosis of AD was established according to the 2024 National Institute on Aging and Alzheimer’s Association (NIA-AA) criteria^2^, requiring both Aβ and tau pathology (Aβ-PET positive, tau-PET positive), with supportive evidence from ¹⁸F-FDG-PET indicating characteristic patterns of cerebral hypometabolism. The NC subjects were required to be negative as indicated by both Aβ and tau PET. All participants underwent total-body tau PET/CT imaging, and multimodal brain PET/MR imaging with Aβ and/or FDG tracers. Demographic and clinical data were collected, including age, sex, education, body mass index (BMI), glycemic status, MMSE, and MoCA scores. Additionally, a separate set of three AD patients underwent full-session dynamic total-body tau PET/CT imaging to characterize the whole-body tracer distribution profile and assess the feasibility of extracting low-frequency physiological fluctuation signals from late plateau-phase acquisitions.

Participants were recruited based on the following inclusion criteria: 1) completion of both total-body tau-PET/CT (^18^F-PI-2620) and brain Aβ-PET/MR (^18^F-Florbetapir) imaging; 2) availability of structural MRI, including 3D T1-weighted and T2-fluid-attenuated inversion recovery (FLAIR) sequences (with optional DTI and rs-fMRI where possible); and 3) a maximum interval of two months between all imaging sessions to ensure biomarker synchronicity. Individuals were excluded if they lacked total-body tau-PET/CT data, presented with poor-quality structural MRI (e.g., significant motion artifacts or processing failures identified during visual quality control), or had a history of intracranial neoplasms. The study protocol was approved by the Institutional Review Board of Renji Hospital, Shanghai Jiao Tong University School of Medicine (Approval No. 2020-004), and written informed consent was obtained from all participants prior to enrollment.

Statistical analyses of demographic characteristics and neuropsychological test scores between ADs and NCs were performed using Python. Continuous variables included age, education, BMI, blood glucose level, total intracranial volume, and MMSE and MoCA scores. Sex was the only categorical variable. The normality of continuous data was assessed using the Shapiro-Wilk test. Variables with a normal distribution were compared using independent samples t-tests, while non-normally distributed variables were analyzed using Mann-Whitney U tests. Categorical variables were summarized as counts and analyzed using the chi-square test for independence. Statistical significance was set at *p* < 0.05.

### PET/CT and MRI acquisition

The ^18^F-PI-2620 radiotracer was synthesized at Renji Hospital using a Sumitomo HM-10 cyclotron (Osaka, Japan). The synthesis process strictly adhered to Good Manufacturing Practice standards for sterile and pyrogen-free production, as well as the FDA’s Radiotracer Guidelines. Quality control measures ensured a radiochemical purity of ≥ 95% and endotoxin levels below 5 EU/mL. Furthermore, independent quality assurance, including batch-specific residual-solvent analysis and sterility testing, was verified by XingImaging, LLC (USA).

Total-body PET/CT imaging was performed using a uEXPLORER scanner (United Imaging Healthcare). Participants received an intravenous injection of 185.0 ± 18.5 MBq (5.0 ± 0.5 mCi) of ^18^F-PI-2620. Following a 60-minute uptake phase, a 10-minute continuous PET acquisition was conducted. To mitigate head motion during the scan, participants were comfortably stabilized using foam padding and subject-specific three-dimensional printed head molds. A concurrent low-dose CT scan was acquired for attenuation correction and anatomical localization, utilizing a tube voltage of 120 kV, a modulated tube current of 50-200 mAs, a 512 × 512 matrix, and a slice thickness of 2.68 mm.

Multimodal MRI data were acquired on a 3.0T PET/MR scanner (uPMR 790, United Imaging Healthcare). The structural MRI protocol included a high-resolution T1-weighted sequence (repetition time [TR] = 7.9 ms, echo time [TE] = 3.0 ms, matrix size = 256 × 256, voxel size = 1.00 × 1.00 × 1.00 mm³), and a T2-FLAIR sequence (TR/TE/inversion time [TI] = 9948/102/2680 ms, flip angle = 150°, matrix size = 256 × 256, voxel size = 1.00 × 1.00 × 2.00 mm³). Resting-state functional MRI was acquired using a gradient-echo EPI sequence (TR/TE = 2000/30 ms, flip angle = 80°, matrix size = 64 × 64, field of view = 230 × 230 mm², voxel size = 3.59 × 3.59 × 3.50 mm³, 38 axial slices, no interslice gap, bandwidth = 1910 Hz/Px, acceleration factor = 2), with 240 volumes collected per run. DTI was performed using a single-shot echo-planar imaging sequence with diffusion gradients (TR/TE = 4699/84 ms, matrix size = 128 × 118, voxel size = 1.95 × 1.95 × 4.0 mm³). The DTI protocol comprised two non-diffusion-weighted images (*b* = 0 s/mm²) and 32 diffusion-weighted images (*b* = 1000 s/mm²).

### Dynamic reconstruction of total-body PET

The acquired list-mode PET data were reconstructed using ordered subset expectation maximization algorithms with three iterations, 20 subsets, a 192 × 192 matrix, a 600 mm FOV, 2.886 mm slice thickness, and a Gaussian post-filter with 3-mm full-width-at-half-maximum (FWHM), incorporating time-of-flight and point-spread-function modeling. All PET images were corrected for radioactive decay, scatter, attenuation and randoms. For the three auxiliary AD patients who underwent full-session dynamic imaging, the 70-min PET acquisitions were reconstructed into 94 frames: 30 × 2 s, 12 × 5 s, 6 × 10 s, 4 × 30 s, 25 × 60 s, 15 × 120 s, and 2 × 300 s. For BON construction in the main cohort, the plateau-phase data (between 60 and 70 min) underwent quasi-dynamic reconstruction into 60 consecutive 10-s frames. This frame duration was selected in accordance with established high-temporal-resolution PET protocols, which demonstrate that 10-s sampling effectively captures rapid physiological fluctuations akin to those observed in myocardial perfusion and vascular dynamics without compromising the signal-to-noise ratio on high-sensitivity total-body systems^56^. Furthermore, this sampling frequency provides a sufficient number of time points (n = 60) to robustly characterize macroscopic late-phase PET signal fluctuations which serve as a surrogate for systemic hemodynamic and physiological synchronization establishing a reliable foundation for subsequent connectivity analysis across the brain and extracranial regions.

### BON construction and analysis

To characterize the temporal coupling of late-phase PET signal fluctuations across the brain and extracerebral organs, we implemented a dynamic total-body PET/CT-based workflow comprising cerebral and extracranial preprocessing (**Supplementary Fig. 2a**) and network construction (**Supplementary Fig. 2b**). For cerebral analysis, T1-weighted MRI data underwent reorientation, bias field correction, and parcellation into ten bilateral cortical regions (frontal, parietal, temporal, occipital, insular) using the uAI Research Portal platform^57^. The resulting labels were rigidly registered to a motion-corrected PET reference image and then propagated to the dynamic PET series. For extracranial analysis, total-body CT images were segmented using the nnUNet-based TotalSegmentator model^58^, then refined to nine representative extracranial regions (kidneys, liver, lungs, colon, spleen, skeleton, heart, spinal cord, and muscles) plus the aorta by excluding non-essential structures, removing unstable outputs, and applying mask erosion to mitigate partial volume effects. Extracranial motion was corrected using nonlinear frame-wise registration, after which CT-derived organ masks were mapped into PET space for TAC extraction. The aorta served as the reference region for normalization^59, 60^ of both cerebral and extracranial TACs. To minimize spurious correlations related to coherent physiological motion and shared low-count fluctuations in the late plateau window (e.g., respiration-related displacement of the liver and spinal cord), motion-related nuisance variance was further regressed out before time-series analysis. Specifically, an expanded set of 28 regressors (including temporal derivatives and squared terms of both six motion parameters and DVARS^61^) was regressed from all regional TACs. Pearson correlations were computed on the residual (confound-reduced) regional time series to form dynamic brain-organ connectivity matrices, which served as the basis for subsequent network and association analyses. Full methodological details are provided in **Supplementary Methods**.

Group differences in BON connectivity were assessed in GRETNA with NBS^62^: edge-wise general linear models (GLMs) including the same covariates were thresholded at *p* < 0.05 to form supra-threshold components, and component significance was determined by permutation to control family-wise error (two-sided, component *p* < 0.05).

### Static PET quantification and tau mediation modeling

To quantitatively assess tau protein accumulation in both cerebral and extracranial regions, we applied SUVR analysis to the static (without dynamic reconstruction) total-body tau PET data of the same subjects (**Supplementary Fig. 3**). For cerebral quantification, high-resolution T1-weighted MRI data were preprocessed and segmented into the same ten bilateral cortical regions along with the cerebellar cortex using a protocol similar to that described above, then rigidly registered to the preprocessed static PET image. The regional SUVRs were calculated as mean uptake in each cortical target divided by mean uptake in the cerebellar cortex, which served as reference given its low and stable tau burden in AD^57,63^. For extracranial quantification, total-body CT images were segmented into the same nine regions together with the aorta using TotalSegmentator^58^, followed by erosion to minimize partial volume effects. Then, these images were nonlinearly aligned to the PET image using the same protocol as used in the dynamic PET pipeline, and the aorta was used as the reference region for normalization in each organ^59, 60^. This dual-referencing approach enabled robust comparison of tau tracer distribution across both brain and extracranial regions. Detailed image segmentation, registration, and normalization procedures are described in **Supplementary Methods**.

Group differences in SUVRs of each brain and extracranial region were tested with GLMs (age, sex, and education were added as covariates) using Python. *P* values were corrected for multiple comparisons across all regions using the Benjamini-Hochberg FDR correction (two-sided, *q* < 0.05). Within the ADs, rank-based partial correlations between FDR-significant SUVRs and cognition (MMSE, MoCA) were computed while controlling for age, sex and education, with FDR correction across tests (*q* < 0.05).

To investigate the relationship between cerebral tau burden, BON, and the cognitive scores, we employed structural equation modeling (SEM) using the *semopy* package in Python to perform mediation analysis. We applied PPCA to extract the main components of both brain-organ connectivity and tau burden. Specifically, we derived the BON-PC from brain-organ connections and the SUVR-PC from the SUVRs, respectively. We first conducted partial Spearman correlation analyses separately for BON-PC and SUVR-PC. Next, we examined the partial correlation between SUVR-PC and cognitive scores (MMSE and MoCA). In the mediation model, BON-PC was used as the predictor, SUVR-PC as the mediator, and cognitive scores (MMSE and MoCA) as the outcomes. Age, sex, and education were included as covariates. The model estimated direct, indirect, and total effects within the SEM framework. To assess the robustness of the mediation effect, we used bootstrapping to derive 95% confidence intervals for the indirect effect. An indirect effect was deemed statistically significant when its bootstrapped 95% confidence interval did not include zero.

### Multimodal MRI assessment and mediation analysis

We developed an integrated multimodal MRI processing pipeline using high-resolution T1-weighted MRI, T2-FLAIR, DTI, and rs-fMRI to derive a comprehensive set of glymphatic indices. These included the CPVF, PVSVF, PVSVF-WM, PVSVF-BG, FW-All, FW-WM, FW-BG, lALPS, rALPS, mALPS, and gBOLD-CSF, detailed **in** Supplementary Materials.

For structural analysis, T1-weighted and T2-FLAIR MRI underwent re-orientation, N4 bias-field correction, and AC-PC alignment, followed by brain tissue segmentation (with special focus on the white matter, basal ganglia, ventricles, total intracranial volume [TIV]) using SynthSeg^64^, choroid plexus segmentation with chp_seg:1.0.1^65^, and PVS segmentation excluding WMH^66^ via SEGCSVD^67^, where the PVS within white matter and basal ganglia were labelled as PVS-WM and PVS-BG, respectively, with all volumes normalized to TIV and quality-controlled by an experienced neuroradiologist.

The DTI data were preprocessed in MRtrix3 with noise and artifact removal, susceptibility distortion and eddy-current correction. FW-All, FW-WM, and FW-BG were estimated using a bi-tensor model within the brain. Crucially, PVS and WMH pixels were excluded from these masks to ensure the specificity of the free-water metrics to the interstitial fluid (ISF) compartment, rather than bulk fluid in enlarged perivascular spaces or pathological edema. Fractional Anisotropy (FA) and diffusivity maps were registered to the ICBM template^68^, and the masks of the projection and association fibers were inverse-transformed to the native space for ALPS computation as the ratio of diffusion along perivascular to non-perivascular directions, yielding lALPS, rALPS, and mALPS.

The gBOLD signals within the gray matter were extracted after standard motion correction and band-pass filtering (0.01-0.1 Hz). In contrast, the CSF signal was extracted from the ventricles after detrending and filtering but without motion correction, in order to preserve the physical displacement information of the CSF that is otherwise removed by spatial realignment^69^. gBOLD-CSF coupling was quantified as the peak negative lag correlation coefficient within a ±10-s lag window.

To examine the link among BON, glymphatic function, and cognitive scores in ADs, we also applied PPCA to the 11 indices to extract the primary and the secondary components representing glymphatic function, yielding GLY-PC1 and GLY-PC2. We tested the association between these components and the BON-PC using partial Spearman correlation, adjusting for age, sex, and education. Consistent with the tau model, mediation analyses were performed using SEM, with BON-PC as the predictor, GLY-PC1 or -PC2 as the mediator, and cognitive scores as outcomes, adjusting for age, sex, and education. All other analyses were the same as above. Additionally, we further examined pairwise partial Spearman correlation between each BON connection and individual glymphatic indices in the ADs, adjusting for age, sex, and education.

### Parallel mediation analysis

To disentangle the independent statistical mediating effects of tau pathology and glymphatic function, we constructed a parallel multiple mediation model within the SEM framework. In this unified model, BON-PC served as the predictor, with both SUVR-PC and GLY-PC1 entered as simultaneous mediators, and cognitive scores as the outcomes. Covariates (age, sex, and education) were adjusted for in all pathways. This approach allowed for the estimation of specific indirect effects associated with each mediator while controlling for the other. All other analyses were the same as above.

### Split-half reliability analysis

To evaluate the robustness and temporal stability of the constructed BON, a split-half reliability analysis was performed. The dynamic PET data (comprising 60 time points) were randomly divided into two independent subsets of 30 time points each, which were then processed through the identical pipeline. For each subset, regional time series were extracted for the 19 pre-defined regions (10 cerebral and 9 extracranial regions). Pairwise Pearson correlations were computed to generate two independent connectivity matrices per participant. Connection-level reliability was quantified using the ICC (3,1) ^70^, calculated edge-wise between the two split matrices across the entire cohort. High group-level ICC values were considered indicative of a reproducible network architecture.

### TAC analysis

To characterize macroscopic total-body temporal dynamics and validate the feasibility of extracting low-frequency physiological fluctuation signals from the late-phase plateau, we applied a dynamic reconstruction protocol to the three AD patients who underwent full-session dynamic imaging. We computed time-resolved SUVRs for the 19 brain and extracranial regions across 94 sequential time points, with the late-phase plateau SUV from the aorta serving as the reference region for all targets. This approach facilitated the generation of group-averaged TACs across these individuals, providing a benchmark for comparing the tracer kinetics within the late-phase windows used for BON construction.

## Data Availability

The data underlying the findings of this study are available from the corresponding author upon reasonable request.

## Acknowledgements

We are grateful to all the participants in this study. This work is partially supported by the National Natural Science Foundation of China (Nos. 62571330, 62131015, U23A20295, and 82572344), Shanghai Pilot Program for Basic Research - Chinese Academy of Science, Shanghai Branch (No. JCYJ-SHFY-2022-014), the Construction Project of Shanghai Key Laboratory of Molecular Imaging (No. 18DZ2260400), Shanghai Aging Women and Children’s Health Research Program (No. 2020YJZX0107), Shanghai Zhangjiang National Innovation Demonstration Zone Special Funds for Major Projects (No. ZJ2018-ZD-012), and Shenzhen Science and Technology Program (No. KCXFZ20211020163408012). The computation in this work was supported by the HPC Platform of ShanghaiTech University.

## Author contributions

**H.Z.** and **C.Z.** conceived the study, designed the research framework, and oversaw the project execution and supervision. **L.W.** performed the neuroimaging data analysis and implemented quality control protocols. **L.L.**, **Yan.Z.**, **M.X.**, **J.L.**, and **C.Z.** were responsible for clinical data acquisition and the interpretation of neuroimaging findings. **L.W.**, **Y.J.**, and **Y.T.** generated the figures and drafted the manuscript. **Y.J.**, **J.Y.**, **Yaj.Z.**, **Y.W.,** and **F.S.** contributed to the critical revision and refinement of the manuscript for intellectual content. All authors reviewed and approved the final version of the manuscript.

## Competing interests

F.S. is an employee of United Imaging Intelligence, Shanghai, China. The company had no role in the design or conduct of the study, or in the analysis and interpretation of the data. All other authors report no conflicts of interest relevant to this article.

## Code availability

Custom code used to generate the results is available from the corresponding authors upon reasonable request.

## Notes

### Author Declarations

Institutional Review Board of Renji Hospital, Shanghai Jiao Tong University School of Medicine gave ethical approval for this work (Approval No. 2020-004).

## References

1. Pereira, J.B., et al. Untangling the association of amyloid-β and tau with synaptic and axonal loss in Alzheimer’s disease. Brain 144, 310–324 (2021).

2. Jack, C.R., Jr., et al. Revised criteria for diagnosis and staging of Alzheimer’s disease: Alzheimer’s Association Workgroup. Alzheimers Dement 20, 5143–5169 (2024).

3. Busche, M.A. & Hyman, B.T. Synergy between amyloid-beta and tau in Alzheimer’s disease. Nat Neurosci 23, 1183–1193 (2020).

4. Ossenkoppele, R., et al. Accuracy of Tau Positron Emission Tomography as a Prognostic Marker in Preclinical and Prodromal Alzheimer Disease: A Head-to-Head Comparison Against Amyloid Positron Emission Tomography and Magnetic Resonance Imaging. JAMA Neurol 78, 961–971 (2021).

5. Zhang, Y., Chen, H., Li, R., Sterling, K. & Song, W. Amyloid β-based therapy for Alzheimer’s disease: challenges, successes and future. Signal Transduct Target Ther 8, 248 (2023).

6. Ye, J., Wan, H., Chen, S. & Liu, G.P. Targeting tau in Alzheimer’s disease: from mechanisms to clinical therapy. Neural Regen Res 19, 1489–1498 (2024).

7. Jorfi, M., Maaser-Hecker, A. & Tanzi, R.E. The neuroimmune axis of Alzheimer’s disease. Genome Med 15, 6 (2023).

8. Li, H., et al. Longitudinal assessment of peripheral organ metabolism and the gut microbiota in an APP/PS1 transgenic mouse model of Alzheimer’s disease. Neural Regen Res 20, 2982–2997 (2025).

9. Waigi, E.W., et al. Soluble and insoluble protein aggregates, endoplasmic reticulum stress, and vascular dysfunction in Alzheimer’s disease and cardiovascular diseases. Geroscience 45, 1411–1438 (2023).

10. Saher, G. Cholesterol Metabolism in Aging and Age-Related Disorders. Annu Rev Neurosci 46, 59–78 (2023).

11. Louveau, A., Da Mesquita, S. & Kipnis, J. Lymphatics in Neurological Disorders: A Neuro-Lympho-Vascular Component of Multiple Sclerosis and Alzheimer’s Disease? Neuron 91, 957–973 (2016).

12. Wu, B., et al. Liver as a new target organ in Alzheimer’s disease: insight from cholesterol metabolism and its role in amyloid-beta clearance. Neural Regen Res 20, 695–714 (2025).

13. Valenza, G., Matić, Z. & Catrambone, V. The brain-heart axis: integrative cooperation of neural, mechanical and biochemical pathways. Nat Rev Cardiol (2025).

14. Hablitz, L.M. & Nedergaard, M. The Glymphatic System: A Novel Component of Fundamental Neurobiology. J Neurosci 41, 7698–7711 (2021).

15. Yin, J., et al. Mechanistic Insights and Emerging Therapeutic Targets of Alzheimer’s Disease: From the Perspective of Inter-Organ Crosstalk. Aging Dis (2024).

16. Lin, K.J., Huang, S.Y., Huang, K.L., Huang, C.C. & Hsiao, I.T. Human biodistribution and radiation dosimetry for the tau tracer [(18)F]Florzolotau in healthy subjects. EJNMMI Radiopharm Chem 9, 27 (2024).

17. Gu, F. & Wu, Q. Quantitation of dynamic total-body PET imaging: recent developments and future perspectives. Eur J Nucl Med Mol Imaging 50, 3538–3557 (2023).

18. Bhattarai, A., et al. Kinetic modeling of (18)F-PI-2620 binding in the brain using an image-derived input function with total-body PET. EJNMMI Res 15, 62 (2025).

19. Reed, M.B., et al. Whole-body metabolic connectivity framework with functional PET. Neuroimage 271, 120030 (2023).

20. Jiang-Xie, L.F., Drieu, A. & Kipnis, J. Waste clearance shapes aging brain health. Neuron 113, 71–81 (2025).

21. Devesa, A., et al. Multiorgan Imaging for Interorgan Crosstalk in Cardiometabolic Diseases. Circ Res 136, 1454–1475 (2025).

22. Ritson, M., Wheeler-Jones, C.P.D. & Stolp, H.B. Endothelial dysfunction in neurodegenerative disease: Is endothelial inflammation an overlooked druggable target? J Neuroimmunol 391, 578363 (2024).

23. Zhu, W.M., Neuhaus, A., Beard, D.J., Sutherland, B.A. & DeLuca, G.C. Neurovascular coupling mechanisms in health and neurovascular uncoupling in Alzheimer’s disease. Brain 145, 2276–2292 (2022).

24. Nair, S.S., et al. Investigation of Autonomic Dysfunction in Alzheimer’s Disease-A Computational Model-Based Approach. Brain Sci 13(2023).

25. Leipp, F., et al. Glial fibrillary acidic protein in Alzheimer’s disease: a narrative review. Brain Commun 6, fcae396 (2024).

26. Moyse, E., et al. Neuroinflammation: A Possible Link Between Chronic Vascular Disorders and Neurodegenerative Diseases. Front Aging Neurosci 14, 827263 (2022).

27. Nakagawa, K., et al. Autoregulation in the posterior circulation is altered by the metabolic state of the visual cortex. Stroke 40, 2062–2067 (2009).

28. Louveau, A., et al. Structural and functional features of central nervous system lymphatic vessels. Nature 523, 337–341 (2015).

29. Larson, A.S., Lanzino, G. & Brinjikji, W. Variations of Intracranial Dural Venous Sinus Diameters from Birth to 20 Years of Age: An MRV-Based Study. AJNR Am J Neuroradiol 41, 2351–2357 (2020).

30. Liu, L., et al. Anatomy imaging and hemodynamics research on the cerebral vein and venous sinus among individuals without cranial sinus and jugular vein diseases. Front Neurosci 16, 999134 (2022).

31. Ottoy, J., et al. Tau follows principal axes of functional and structural brain organization in Alzheimer’s disease. Nat Commun 15, 5031 (2024).

32. Nagai, M., Hoshide, S. & Kario, K. The insular cortex and cardiovascular system: a new insight into the brain-heart axis. J Am Soc Hypertens 4, 174–182 (2010).

33. Tu, Z., et al. Tauopathy promotes spinal cord-dependent production of toxic amyloid-beta in transgenic monkeys. Signal Transduct Target Ther 8, 358 (2023).

34. Wu, Z., et al. Spinal cord injury-activated C/EBPβ-AEP axis mediates cognitive impairment through APP C586/Tau N368 fragments spreading. Prog Neurobiol 227, 102467 (2023).

35. Wang, J., Gu, B.J., Masters, C.L. & Wang, Y.J. A systemic view of Alzheimer disease - insights from amyloid-beta metabolism beyond the brain. Nat Rev Neurol 13, 703 (2017).

36. Cheng, Y., et al. Physiological beta-amyloid clearance by the liver and its therapeutic potential for Alzheimer’s disease. Acta Neuropathol 145, 717–731 (2023).

37. Wang, S., et al. Association of impaired kidney function with dementia and brain pathologies: A community-based cohort study. Alzheimers Dement 19, 2765–2773 (2023).

38. Elia, A. & Fossati, S. Autonomic nervous system and cardiac neuro-signaling pathway modulation in cardiovascular disorders and Alzheimer’s disease. Front Physiol 14, 1060666 (2023).

39. Roy, B., et al. Reduced regional cerebral blood flow in patients with heart failure. Eur J Heart Fail 19, 1294–1302 (2017).

40. Xie, Z., Tong, S., Chu, X., Feng, T. & Geng, M. Chronic Kidney Disease and Cognitive Impairment: The Kidney-Brain Axis. Kidney Dis (Basel) 8, 275–285 (2022).

41. Yang, C., et al. Single-cell transcriptomics identifies premature aging features of TERC-deficient mouse brain and bone marrow. Geroscience 44, 2139–2155 (2022).

42. Astarita, G., et al. Deficient liver biosynthesis of docosahexaenoic acid correlates with cognitive impairment in Alzheimer’s disease. PLoS One 5, e12538 (2010).

43. La Joie, R., et al. Prospective longitudinal atrophy in Alzheimer’s disease correlates with the intensity and topography of baseline tau-PET. Sci Transl Med 12(2020).

44. Nedergaard, M. & Goldman, S.A. Glymphatic failure as a final common pathway to dementia. Science 370, 50–56 (2020).

45. Lipsitz, L.A. & Goldberger, A.L. Loss of ’complexity’ and aging. Potential applications of fractals and chaos theory to senescence. JAMA 267, 1806–1809 (1992).

46. Iadecola, C. The Neurovascular Unit Coming of Age: A Journey through Neurovascular Coupling in Health and Disease. Neuron 96, 17–42 (2017).

47. Iturria-Medina, Y., et al. Early role of vascular dysregulation on late-onset Alzheimer’s disease based on multifactorial data-driven analysis. Nat Commun 7, 11934 (2016).

48. Zlokovic, B.V. Neurovascular pathways to neurodegeneration in Alzheimer’s disease and other disorders. Nat Rev Neurosci 12, 723–738 (2011).

49. Wu, J.W., et al. Neuronal activity enhances tau propagation and tau pathology in vivo. Nat Neurosci 19, 1085–1092 (2016).

50. Mestre, H., et al. Flow of cerebrospinal fluid is driven by arterial pulsations and is reduced in hypertension. Nat Commun 9, 4878 (2018).

51. Benveniste, H., et al. The Glymphatic System and Waste Clearance with Brain Aging: A Review. Gerontology 65, 106–119 (2019).

52. Dumont, M., et al. Free Water in White Matter Differentiates MCI and AD From Control Subjects. Front Aging Neurosci 11, 270 (2019).

53. Fultz, N.E., et al. Coupled electrophysiological, hemodynamic, and cerebrospinal fluid oscillations in human sleep. Science 366, 628–631 (2019).

54. Badawi, R.D., et al. First Human Imaging Studies with the EXPLORER Total-Body PET Scanner. J Nucl Med 60, 299–303 (2019).

55. Rahmim, A., et al. Dynamic whole-body PET imaging: principles, potentials and applications. Eur J Nucl Med Mol Imaging 46, 501–518 (2019).

56. Otaki, Y., et al. Improved myocardial blood flow estimation with residual activity correction and motion correction in (18)F-flurpiridaz PET myocardial perfusion imaging. Eur J Nucl Med Mol Imaging 49, 1881–1893 (2022).

57. Wu, J., et al. uRP: An integrated research platform for one-stop analysis of medical images. Front Radiol 3, 1153784 (2023).

58. Wasserthal, J., et al. TotalSegmentator: Robust Segmentation of 104 Anatomic Structures in CT Images. Radiol Artif Intell 5, e230024 (2023).

59. Dubroff, J.G., et al. [(11)C]Carfentanil PET Whole-Body Imaging of μ-Opioid Receptors: A First in-Human Study. J Nucl Med (2025).

60. Holy, E.N., et al. Non-invasive kinetic modeling of [18F]-florbetaben and [18F]-PI-2620 with total-body dynamic EXPLORER PET. Alzheimer’s & Dementia 19, e075087 (2023).

61. Afyouni, S. & Nichols, T.E. Insight and inference for DVARS. Neuroimage 172, 291–312 (2018).

62. Wang, J., et al. GRETNA: a graph theoretical network analysis toolbox for imaging connectomics. Front Hum Neurosci 9, 386 (2015).

63. Groot, C., Villeneuve, S., Smith, R., Hansson, O. & Ossenkoppele, R. Tau PET Imaging in Neurodegenerative Disorders. J Nucl Med 63, 20s–26s (2022).

64. Billot, B., et al. SynthSeg: Segmentation of brain MRI scans of any contrast and resolution without retraining. Med Image Anal 86, 102789 (2023).

65. Eisma, J.J., et al. Deep learning segmentation of the choroid plexus from structural magnetic resonance imaging (MRI): validation and normative ranges across the adult lifespan. Fluids Barriers CNS 21, 21 (2024).

66. Mojiri Forooshani, P., et al. Deep Bayesian networks for uncertainty estimation and adversarial resistance of white matter hyperintensity segmentation. Hum Brain Mapp 43, 2089–2108 (2022).

67. Gibson, E., et al. segcsvdWMH: A Convolutional Neural Network-Based Tool for Quantifying White Matter Hyperintensities in Heterogeneous Patient Cohorts. (Wiley Online Library, 2024).

68. Mori, S., et al. Stereotaxic white matter atlas based on diffusion tensor imaging in an ICBM template. Neuroimage 40, 570–582 (2008).

69. Jiang, D., et al. Regional Glymphatic Abnormality in Behavioral Variant Frontotemporal Dementia. Ann Neurol 94, 442–456 (2023).

70. Chen, G., et al. Intraclass correlation: Improved modeling approaches and applications for neuroimaging. Hum Brain Mapp 39, 1187–1206 (2018).

