## Supplementary for "Brain-Organ Hypersynchrony and Cognitive Decline in Alzheimer’s Disease: Potential Links with Tauopathy and Glymphatic Dysfunction"

### **Supplementary** **Figures**

#### **Supplementary Figure 1**

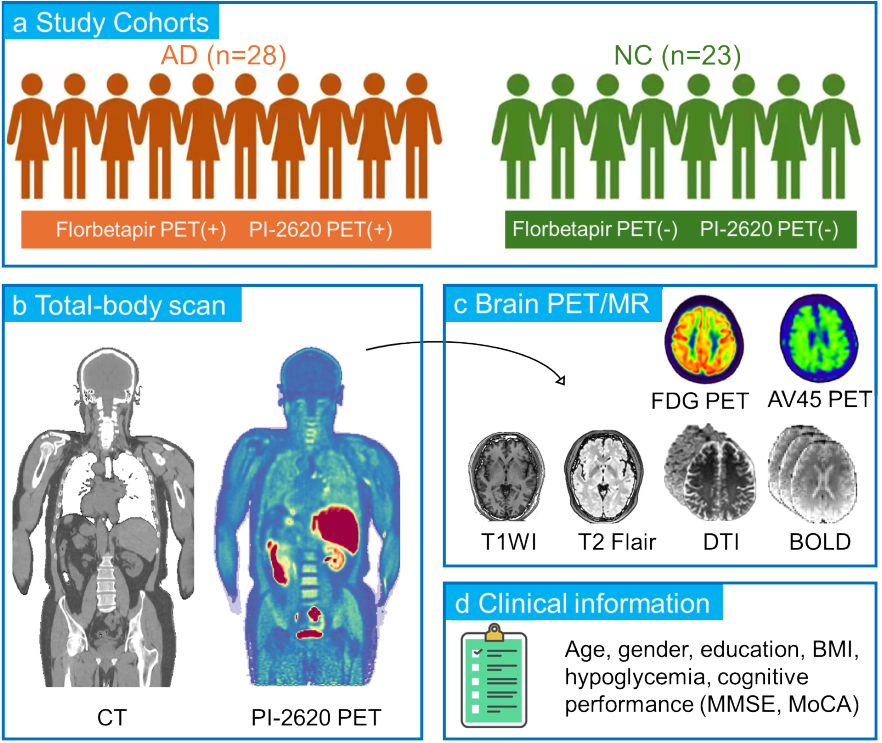

**Supplementary Fig. 1 |** **Overview of study cohorts, imaging acquisitions, and collected clinical information. a** Study cohorts included 28 patients with Alzheimer’s disease (AD; positive for both ^18^F-florbetapir and ^18^F-PI-2620 PET) and 23 age-, sex-, and education-matched negative controls (NC; negative for both PET tracers). **b** Total-body PET/CT acquisitions consisted of structural CT (left) and ^18^F-PI-2620 PET (right) scans. **c** Brain imaging included multimodal PET/MR scans: ^18^F-FDG PET, ^18^F-AV45 PET, T1-weighted MRI, T2-FLAIR, DTI, and rs-fMRI. **d** Clinical and demographic information collected included age, sex, education, BMI, glycemic status, and cognitive performance assessed with MMSE and MoCA.

*Abbreviations:* AD, Alzheimer’s disease; NC, negative controls; PET, positron emission tomography; MRI, magnetic resonance imaging; DTI, diffusion tensor imaging; BOLD, blood-oxygen-level-dependent; BMI, body mass index; MMSE, Mini-Mental State Examination; MoCA, Montreal Cognitive Assessment.

#### **Supplementary Figure 2**

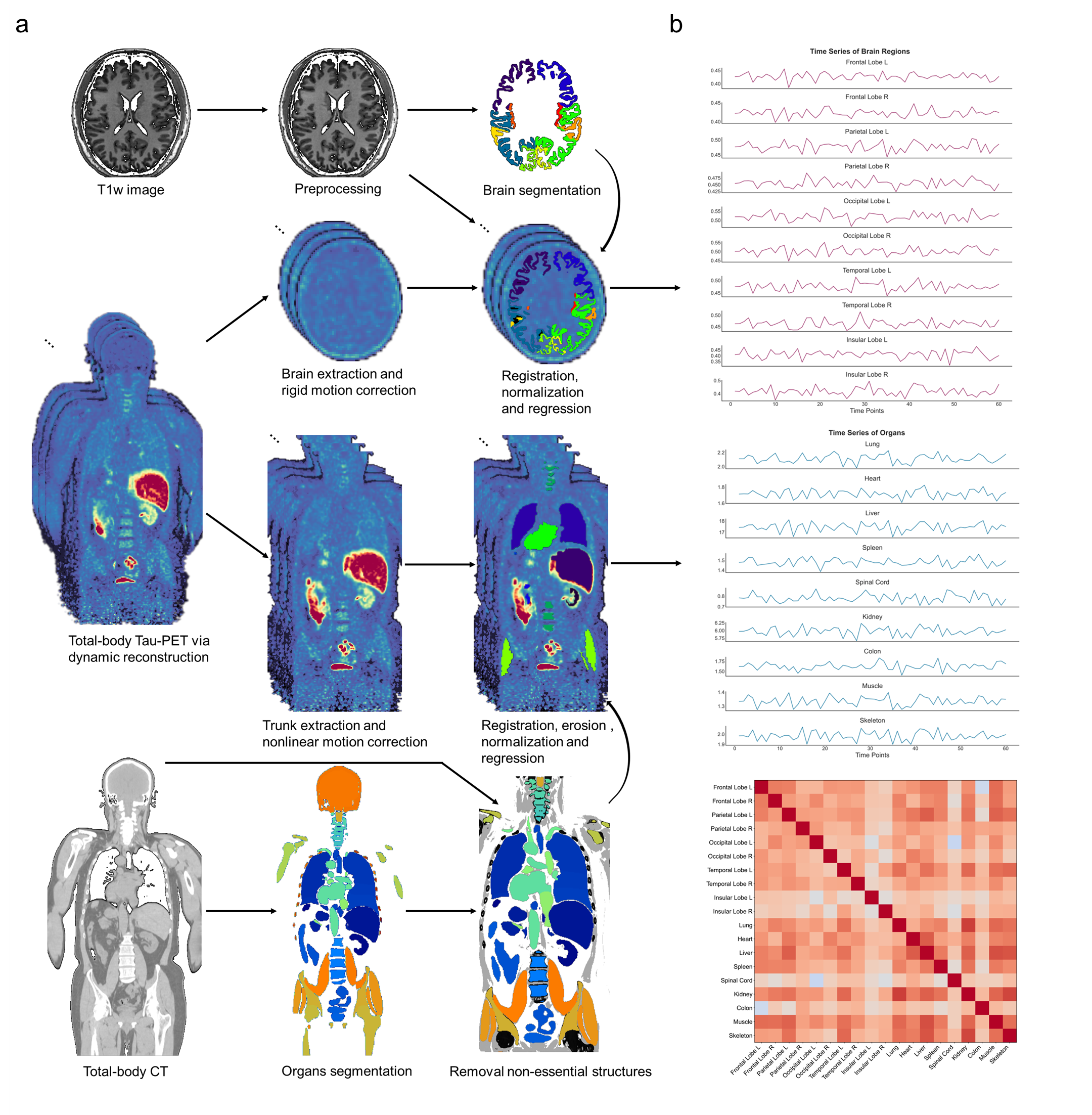

**Supplementary Fig. 2 | Dynamic total-body PET framework for brain-organ network construction. a** Workflow of dynamic total-body tau-PET analysis integrating brain MRI and total-body CT. Top: T1-weighted MRI preprocessing and brain segmentation into cortical regions. Middle: Dynamic total-body PET reconstruction, brain and trunk extraction, motion correction, registration, normalization, regression, and related preprocessing steps. Bottom: CT-based segmentation of major organs, removal of non-essential structures. **b** Representative dynamic brain-organ connectivity matrix derived from correlations between regional time series.

#### **Supplementary Figure 3**

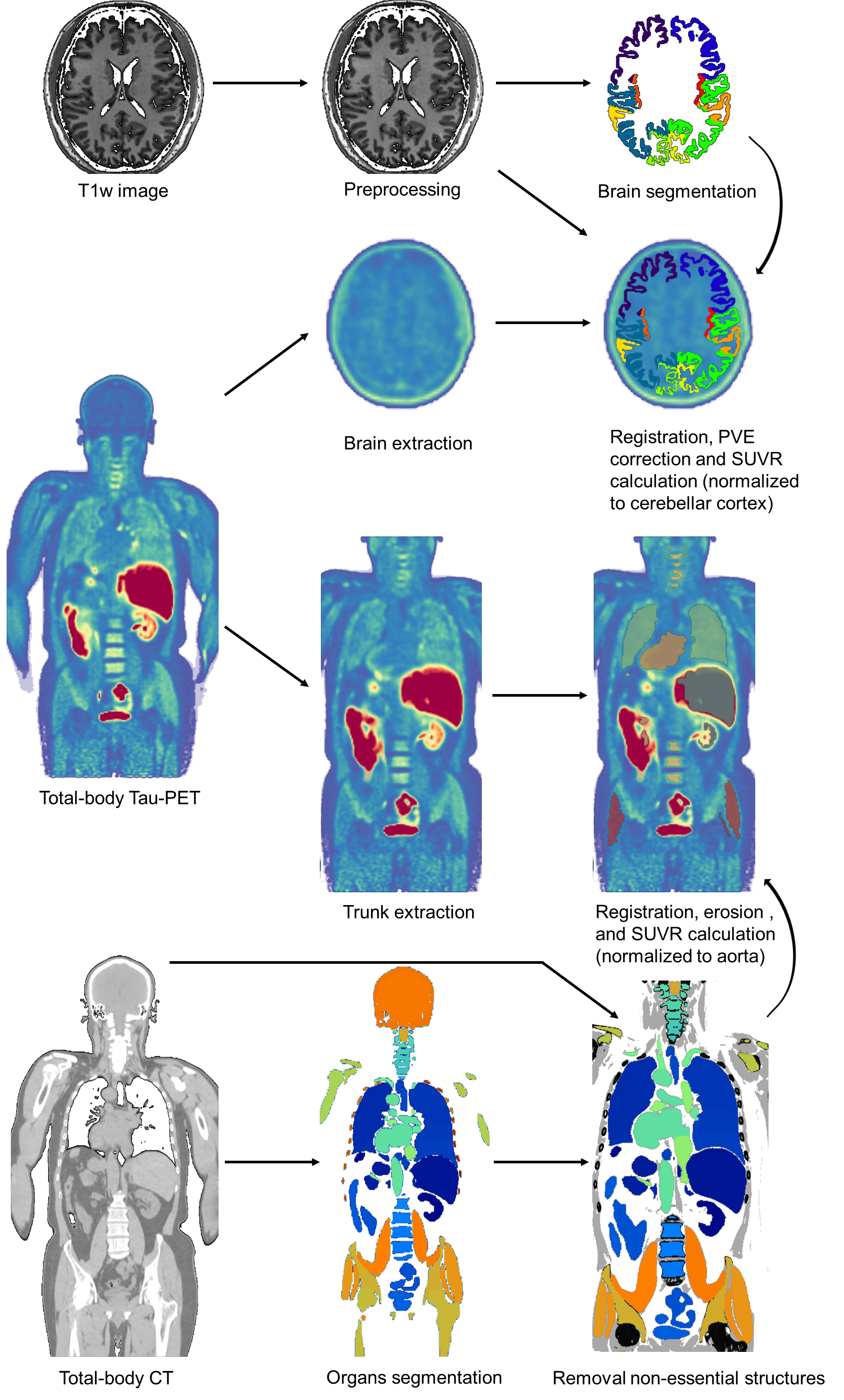

**Supplementary Fig. 3 | Static PET analysis workflow for cerebral and extracranial SUVR quantification.** Workflow of static total-body tau-PET analysis integrating high-resolution brain MRI and total-body CT. Top: T1-weighted MRI preprocessing and cortical segmentation. Middle: Brain and Trunk extraction, registration, normalization, SUVR calculation, and related preprocessing steps. Bottom: CT-based organ segmentation, and removal of non-essential structures.

#### **Supplementary Figure 4**

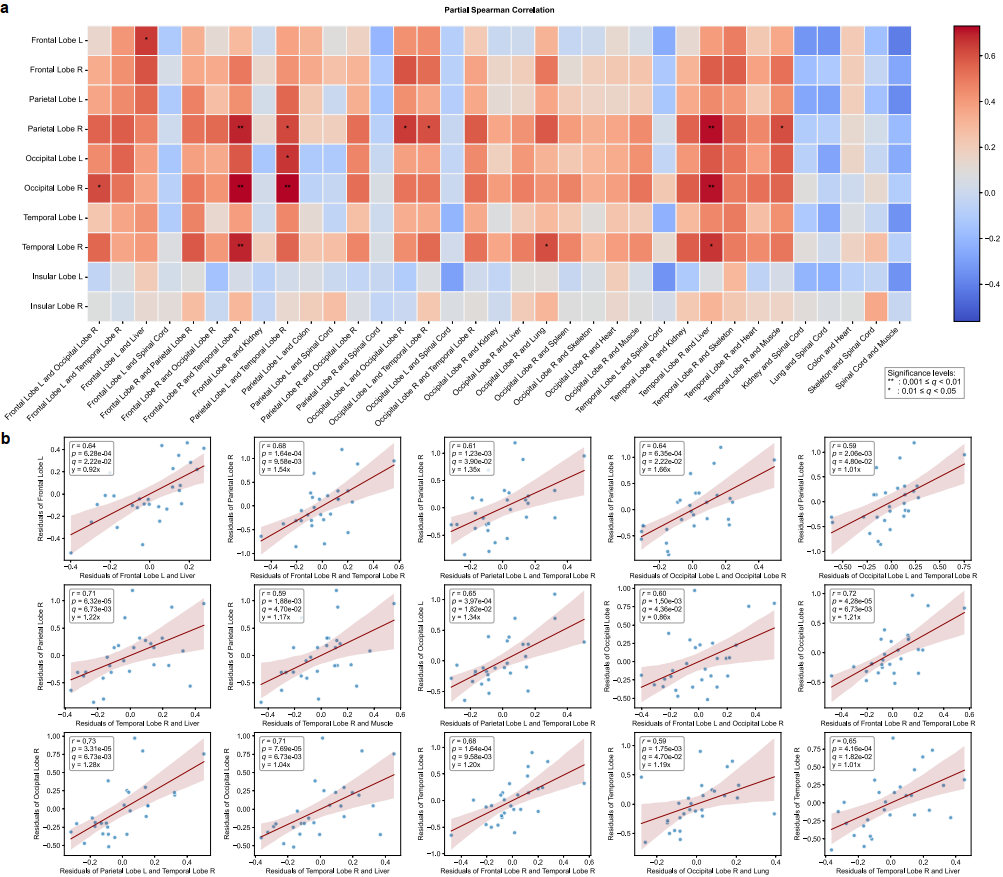

**Supplementary Fig. 4 | Partial Spearman correlations between brain-organ connectivity and regional tau SUVRs in AD. a** Heatmap showing partial Spearman correlation coefficients between brain-organ connections and cerebral tau SUVRs, adjusted for age, sex, and education. Statistical significance was determined using false discovery rate (FDR) correction (**q* < 0.05; ***q* < 0.01). The color scale denotes correlation strength, with darker red indicating stronger positive correlations and blue indicating negative correlations. **b** Scatter plots for the 15 significant associations identified in panel a. Each plot shows the adjusted partial Spearman correlation with a fitted least-squares regression line. *r* indicates the correlation coefficient; *p*, the uncorrected *P* value; and *q*, the FDR-adjusted *q* value (*q* < 0.05).

*Abbreviations:* AD, Alzheimer’s disease.

#### **Supplementary Figure 5**

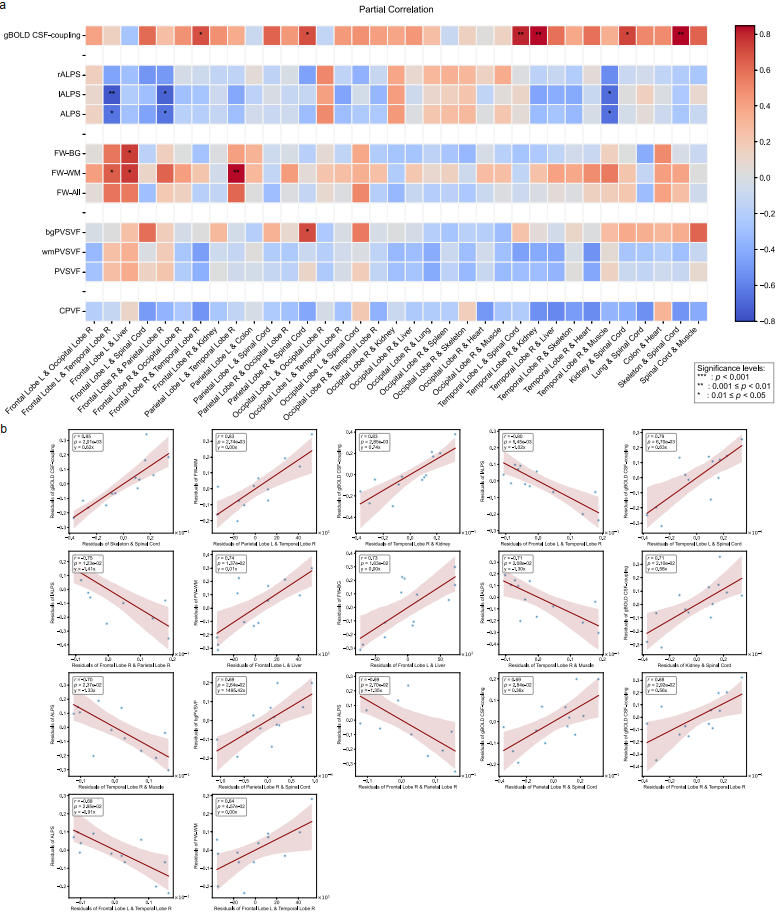

**Supplementary Fig. 5 | Partial Spearman correlations between brain-organ connectivity and glymphatic indices in AD. a** Heatmap showing partial Spearman correlation coefficients between brain-organ connections and glymphatic indices, adjusted for age, sex, and education. **b** Scatter plots for the 17 significant associations identified in panel a. Each plot shows the adjusted partial Spearman correlation with a fitted least-squares regression line.

#### **Supplementary Figure 6**

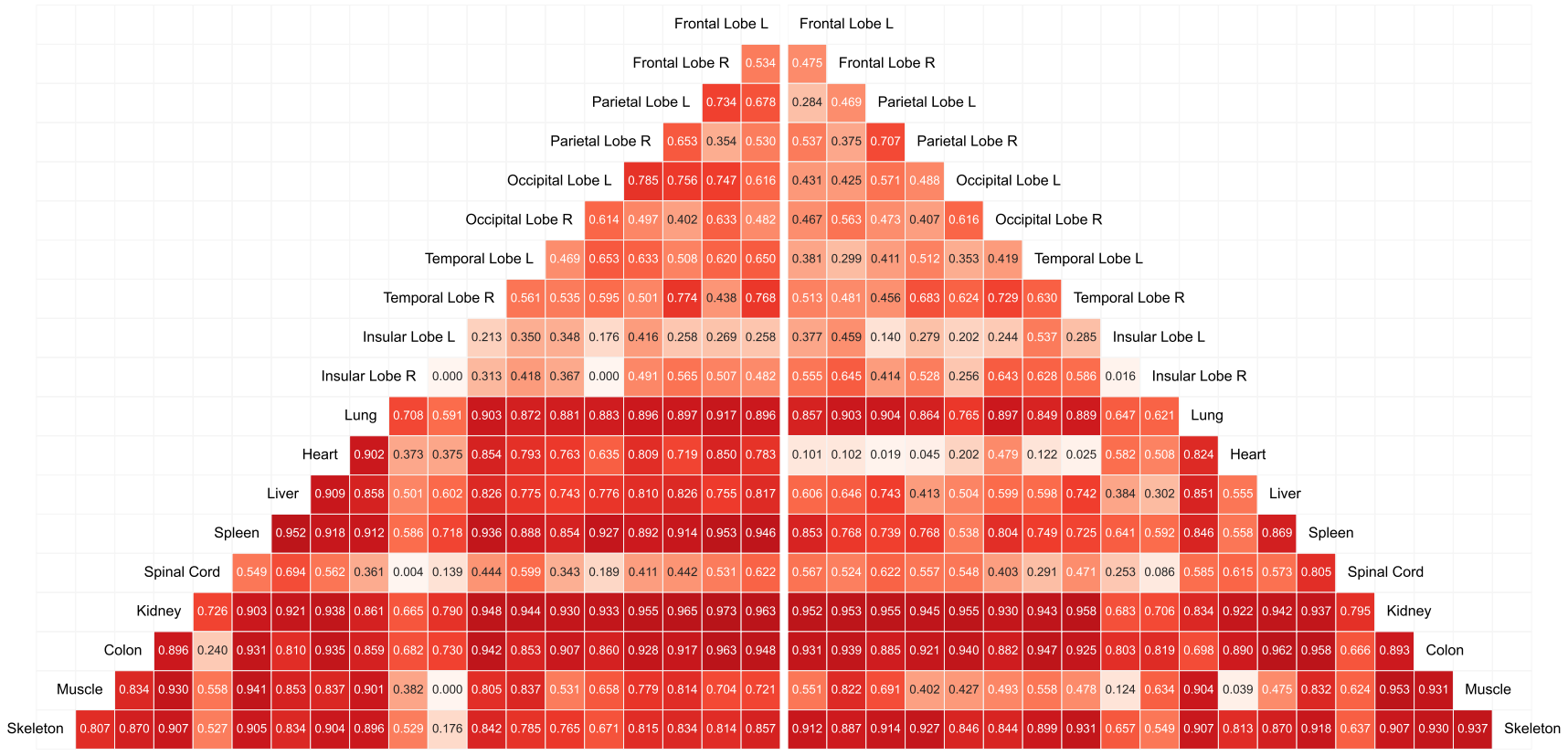
 **Supplementary Fig. 6 | The ICC analysis results of brain-organ connectivity.** Split-half ICC analyses were performed to evaluate the reproducibility of BON in AD (left) and NC (right). The heatmap displays ICC values for all pairwise brain-organ connections, with warmer colors indicating higher reproducibility. Approximately 85% of connections showed ICC values exceeding 0.4, confirming satisfactory measurement stability.

#### **Supplementary Figure 7**

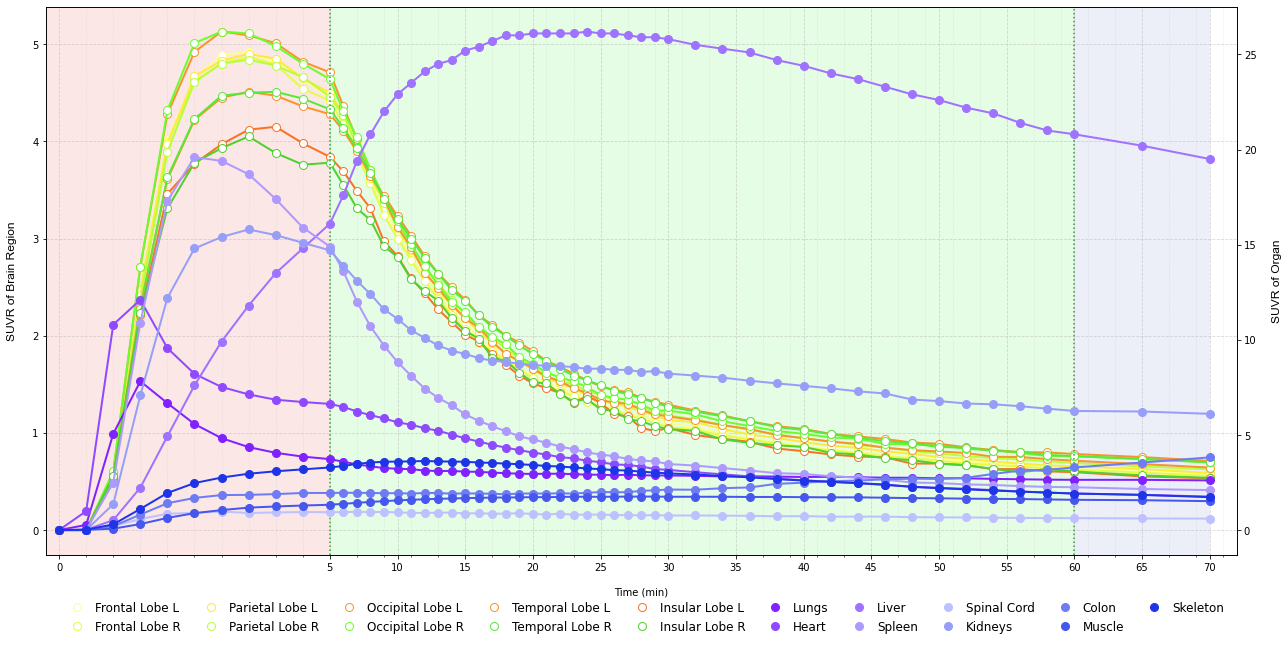

**Supplementary Fig. 7 | Time**-**activity curves of tracer uptake across brain regions and extracranial organs.** TACs are expressed as SUVRs using the late-phase plateau SUV from the aorta serving as the reference region. Representative curves are shown for major cerebral regions (frontal, parietal, occipital, temporal, and insular lobes) and extracranial organs (lungs, heart, liver, spleen, spinal cord, kidneys, colon, muscle, and skeleton). The curves illustrate the temporal ordering of tracer uptake across tissues and the approach to a pseudo-steady state by approximately 60–70 min post-injection.

*Abbreviations:* SUVRs, standardized uptake value ratios; TACs, time-activity curves.

### **Supplementary Methods**

#### **The Analysis Methods of PET via Dynamic Reconstruction**

To characterize the spatiotemporal dynamics of tau tracer uptake across the cerebral and extracranial regions, we established a dynamic total-body PET analysis workflow that integrates high-resolution structural brain MRI and total-body CT for anatomical segmentation and PET-space registration comprising cerebral preprocessing, extracranial preprocessing and BON construction (Supplementary Fig. 2).

For cerebral analysis, T1-weighted MRI images were initially realigned to a standardized orientation, followed by N4 bias field correction^1^ to mitigate intensity non-uniformity caused by magnetic field inhomogeneities. For brain parenchymal segmentation, the mid-scale brain segmentation model available on the uAI Research Portal (United Imaging Healthcare) was used to delineate ten major cortical brain regions (the bilateral frontal, parietal, occipital, temporal, and insular lobes). Dynamic PET volumes were cropped to the brain parenchyma to remove extracranial structures, followed by frame-wise head motion correction. This yielded a motion-corrected mean PET image and six motion parameters (three translations, three rotations). Individual T1-weighted MRI scans were registered to the motion-corrected mean PET image using rigid-body registration in SPM, ensuring precise anatomical alignment. The rigid transforms were then used to map the pre-segmented cortical regions (from the uAI Research Portal) into PET space.

For extracranial analysis, total-body CT images were segmented using the state-of-the-art nnUNet-based TotalSegmentator model^2^, which provides automated segmentation of 116 anatomical structures. Post-segmentation, non-essential structures, such as the skull and limbs, were computationally removed, and unstable segmentation outputs were excluded. Functionally similar structures were merged to establish a refined set of nine representative extracranial regions including the kidneys, liver, lungs, colon, spleen, skeleton, heart, spinal cord, and muscles (skeletal muscles) plus aorta for analysis. This process provides robust anatomical information for precise regional quantification^3^. The dynamic total-body PET images were cropped to exclude the skull and limbs, retaining the trunk. To correct for respiratory and other physiological motion, nonlinear frame-wise registration was performed using the Symmetric Normalization (SyN) algorithm in ANTs. Motion stability was evaluated using the DVARS metric, calculated as the root mean square of voxel-wise intensity differences between consecutive frames^4^. The co-acquired CT image was nonlinearly registered to the motion-corrected mean PET, and representative regions (kidneys, liver, lungs, colon, spleen, skeleton, heart, spinal cord, and muscles) plus the aorta generated based on the TotalSegmentator model were aligned to PET space^2^. A subsequent erosion algorithm was applied to these masks to mitigate partial volume effects and ensure inclusion of only the core, most representative tissue of each organ.

For each dynamic frame, SUVR images were computed using the aorta as the reference region for both cerebral and extracranial targets to ensure a unified scale for total-body interaction analysis ^5, 6^. To rigorously account for residual motion, expanded motion regressors were derived prior to TAC extraction. For head motion, the six original parameters were augmented with their temporal derivatives, squared terms, and squared derivatives (24 regressors total). For organ motion, DVARS was similarly expanded to four regressors. In total, 28 motion-related variables were regressed from the dynamic TACs. This rigorous motion correction procedure ensured the reliability and interpretability of subsequent brain-organ network (BON) analyses. Following comprehensive motion correction, dynamic time series of SUVRs were extracted for each of the ten brain regions and nine extracranial regions across all the available time points. The Pearson correlation coefficient was then calculated between the time series of every pair of regions (brain-brain, organ-organ, and brain-organ) to quantify their statistical dependencies. This pairwise correlation matrix formed the basis for constructing BON, in which nodes represent individual cerebral or extracranial regions, and edge weights reflect the strength of their dynamic functional connectivity. These networks were used for subsequent statistical and clinical association analyses.

#### **Static PET Analysis Methods**

To quantify tau protein accumulation in both cerebral and extracranial regions, we applied a standardized uptake value ratio (SUVR) analysis to static total-body PET data. Image processing followed a multi-step pipeline, integrating high-resolution structural brain MRI and total-body CT to enable precise anatomical segmentation and registration. All procedures were conducted in PET space to ensure spatial consistency (Supplementary Fig. 3).

To calculate the SUVRs for all the brain regions, high-resolution T1-weighted MRI data underwent orientation, bias field correction, and segmentation into ten cortical regions and the cerebellar cortex using the uAI Research Portal. The cerebellar cortex was specifically chosen as the reference region for subsequent PET analysis because of its relatively low and stable tau accumulation in typical AD pathology^7,8^. Total-body PET images were cropped to extract the head volume, followed by partial volume effect (PVE) correction to mitigate spill-over between adjacent regions. Subsequently, the rigid registration algorithm within the Statistical Parametric Mapping (SPM) software was applied to register the pre-processed T1-weighted MRI images to the corresponding PET images. This ensured accurate spatial alignment between the anatomical structures and functional tracer uptake data of the participants. During this registration step, the previously segmented ten brain regions and the cerebellar cortex, derived from the uAI Research Portal platform, were precisely mapped to the PET space. The cerebellar cortex served as the reference region for the intensity normalization of the PET images. The SUVR for each of the ten brain regions was calculated by dividing the mean tracer uptake within each region by the mean uptake in the cerebellar cortex.

Consistent with the dynamic pipeline, extracranial regions were derived from total-body CT using the nnUNet–based TotalSegmentator (116 labels) and post-processed to remove non-essential/unstable labels and merge functionally similar tissues, yielding nine representative regions plus aorta. PET volumes were cropped to exclude skull and limbs to focus on the torso. The corresponding CT was nonlinearly registered to native PET space, and the resulting segmentations were mapped to PET to provide robust anatomical priors for regional quantification. To mitigate partial volume effects, a subsequent erosion algorithm was applied to these masks, ensuring that only the core, most representative tissue of each organ was included in the quantification. The aorta was selected as the reference region for extracranial regions normalization^5, 6^. SUVRs for the nine major extracranial regions (kidneys, liver, heart, lungs, colon, spleen, skeleton, spinal cord, and muscles) were then calculated by normalizing their mean tracer uptake to the mean uptake in the aortic reference region.

This comprehensive approach yielded a robust set of SUVRs for ten major brain regions and nine representative extracranial regions, forming the basis for subsequent statistical analyses of static tau accumulation patterns across the body.

#### **Evaluation of the glymphatic system**

Multimodal MRI comprised high-resolution T1-weighted and T2-FLAIR anatomical scans, diffusion tensor imaging (DTI) and resting-state fMRI (rs-fMRI). These data were used to derive a comprehensive set of glymphatic indices: choroid plexus volume fraction (CPVF), perivascular-space volume fractions (PVSVF; white-matter and basal-ganglia components PVSVF-WM and PVSVF-BG), free-water (FW-All, FW-WM, FW-BG), ALPS indices (left, right and mean; lALPS, rALPS, mALPS) and global BOLD-CSF coupling (gBOLD-CSF). The detailed workflow is summarized below.

T1 and T2-FLAIR preprocessing included reorientation, N4 bias-field correction and AC-PC alignment. Brain tissue segmentation (white matter, basal ganglia, fourth ventricle and total intracranial volume [TIV]) was performed with SynthSeg^9,10^. Choroid plexus (CP) segmentation used the deep learning model chp_seg:1.0.1 (<https://github.com/Center-of-Imaging-Biomarker-Development/chp_seg>) which provides separate results for the left and right hemispheres^11^. CP volumes were normalized to TIV to obtain CPVF. To segment WMH from T2-FLAIR images, we utilized WMH-SynthSeg^12^. Perivascular spaces (PVS) were segmented using SEGCSVD on the basis of the preprocessed T1 and WMH maps^13^, and PVS within white matter and basal ganglia were labelled PVS-WM and PVS-BG, respectively^14-16^; resulting PVS volumes were normalized to TIV to yield PVSVF, PVSVF-WM and PVSVF-BG. All segmentations underwent visual quality control by an experienced neuroradiologist.

The preprocessing of DTI data based on MRtrix3 software (https://www.mrtrix.org) ^17^ involves several crucial steps to ensure data quality^18^. These steps include noise reduction^19^, removal of Gibbs ringing artifacts^20^, susceptibility correction, and eddy current correction^21^. Free-water was estimated using a bi-tensor free-water model implemented in the DiffusionTensorImaging toolbox (https://github.com/sameerd/DiffusionTensorImaging), which implements the bi-tensor diffusion model as originally described by Pasternak et al. ^22^. To quantify free-water content specifically within the brain, white matter and basal ganglia, we constructed a whole brain mask, a white-matter mask and a basal ganglia mask constructed after excluding PVS and WMH, followed by alignment to diffusion space and extraction of voxel-wise free-water values, resulting in the measurement of free-water in whole brain (FW-All), free-water in white matter (FW-WM), free-water in basal ganglia (FW-BG). Subsequently, the fractional anisotropy (FA) and diffusivity maps were generated through the application of the DTIFIT tool provided by FSL (FMRIB Software Library). FA maps were rigidly registered (six degrees of freedom) to the ICBM FA template using FSL FLIRT; the same transforms were applied to the tensor-derived components with appropriate tensor reorientation to preserve principal diffusion directions. The reoriented map included x-axis (reoriented right-left; ro-Dxx), y-axis (reoriented anterior-posterior; ro-Dyy), and z-axis (reoriented inferior-superior; ro-Dzz) directions, along with a reoriented color-coded map^18,23^. Projection and association fibers were selected based on JHU DTI-based white matter atlases (https://identifiers.org/neurovault.image:1408), with corticofugal corona radiata tract fibers representing the projection fibers and superior longitudinal fasciculus representing the association fibers^24,25^. ROI center coordinates in MNI space were at (24, −12, 24) and (−28, −12, 24) for the left/right projection ROIs, and (36, −12, 24) and (−40, −12, 24) for the left/right association ROIs^16,25^, respectively. For each center, a spherical ROI with a 5-mm radius was defined to select fibers running along the body of the lateral ventricle^25,26^. Subsequently, FA and diffusivity maps were nonlinearly registered to the ICBM template using ANTs to obtain the forward and inverse transforms; ROIs were inverse-transformed to native diffusion space to ensure anatomically accurate placement^27,28^. Evaluation of ALPS-index was accordant with previous studies^18,24^. This index is calculated using the following formula: *ALPS index = mean (ro-Dxx, proj, ro-Dxx, assoc) ∕ mean (ro-Dyy, proj, ro-Dzz, assoc)*. ROIs were selected in bilateral hemispheres of the brain, and the lALPS and rALPS indices were calculated for each hemisphere, respectively. Subsequently, the mALPS index was obtained by averaging the left and right ALPS indices, providing a more comprehensive evaluation of glymphatic system function^29,30^.

Rs-fMRI data were preprocessed using the Statistical Parametric Mapping 12 (SPM12) based on MATLAB, following established pipelines (<https://github.com/evavanheese/BOLD-CSF>). Specifically, we performed motion correction, skull stripping, linear trend removal, and temporal filtering (band-pass filter, 0.01-0.1 Hz) ^31,32^. For analysis of the global brain BOLD (gBOLD) signal, rs-fMRI signals within each individual’s gray matter mask were extracted and Z-score normalized at each voxel prior to averaging, ensuring equal fluctuation amplitude across voxels. The average amplitude of these normalized signals, representing the gBOLD signal, reflected the level of global synchronization. As in previous studies, the first 5 and last 5 rs-fMRI volumes were discarded to allow magnetization to reach a steady state and to avoid edge effects from temporal filtering^31,33^. For the extraction of the CSF signal, rs-fMRI data underwent linear trends detrending and band-pass filtering (0.01–0.1 Hz), but without motion correction^34^. To avoid spatial blurring, all rs-fMRI signal analyses were performed in the original spatial frame without transforming into a standard space^34^. We did not perform a nuisance regression analysis for the gBOLD and CSF signal, because these variables were the focus of this study^35^. Spatial smoothing was also omitted since it averages the signal between neighboring voxels, thus degrading spatial resolution of the detection of original pulsation signals ^36^. The time lag corresponding to the maximum negative correlation between the gBOLD and CSF signals within the ± 10 s window was defined as the peak negative lag, and its correlation coefficient was used to quantify BOLD-CSF coupling, consistent with previous studies^31,33^.

### **References**

1.	Tustison, N.J.*, et al.* N4ITK: improved N3 bias correction. *IEEE Trans Med Imaging* **29**, 1310-1320 (2010).

2.	Wasserthal, J.*, et al.* TotalSegmentator: Robust Segmentation of 104 Anatomic Structures in CT Images. *Radiol Artif Intell* **5**, e230024 (2023).

3.	Tian, Y.E.*, et al.* Heterogeneous aging across multiple organ systems and prediction of chronic disease and mortality. *Nat Med* **29**, 1221-1231 (2023).

4.	Afyouni, S. & Nichols, T.E. Insight and inference for DVARS. *Neuroimage* **172**, 291-312 (2018).

5.	Dubroff, J.G.*, et al.* [(11)C]Carfentanil PET Whole-Body Imaging of μ-Opioid Receptors: A First in-Human Study. *J Nucl Med* (2025).

6.	Holy, E.N.*, et al.* Non‐invasive kinetic modeling of [18F]‐florbetaben and [18F]‐PI‐2620 with total‐body dynamic EXPLORER PET. *Alzheimer's & Dementia* **19**, e075087 (2023).

7.	Wu, J.*, et al.* uRP: An integrated research platform for one-stop analysis of medical images. *Front Radiol* **3**, 1153784 (2023).

8.	Landau, S.M.*, et al.* Quantification of amyloid beta and tau PET without a structural MRI. *Alzheimers Dement* **19**, 444-455 (2023).

9.	Billot, B.*, et al.* SynthSeg: Segmentation of brain MRI scans of any contrast and resolution without retraining. *Med Image Anal* **86**, 102789 (2023).

10.	Billot, B.*, et al.* Robust machine learning segmentation for large-scale analysis of heterogeneous clinical brain MRI datasets. *Proc Natl Acad Sci U S A* **120**, e2216399120 (2023).

11.	Eisma, J.J.*, et al.* Deep learning segmentation of the choroid plexus from structural magnetic resonance imaging (MRI): validation and normative ranges across the adult lifespan. *Fluids Barriers CNS* **21**, 21 (2024).

12.	Mojiri Forooshani, P.*, et al.* Deep Bayesian networks for uncertainty estimation and adversarial resistance of white matter hyperintensity segmentation. *Hum Brain Mapp* **43**, 2089-2108 (2022).

13.	Gibson, E.*, et al.* segcsvdWMH: A Convolutional Neural Network‐Based Tool for Quantifying White Matter Hyperintensities in Heterogeneous Patient Cohorts. (Wiley Online Library, 2024).

14.	Kim, J.Y., Nam, Y., Kim, S., Shin, N.Y. & Kim, H.G. MRI-visible Perivascular Spaces in the Neonatal Brain. *Radiology* **307**, e221314 (2023).

15.	Björnfot, C.*, et al.* Cerebral arterial stiffness is linked to white matter hyperintensities and perivascular spaces in older adults - A 4D flow MRI study. *J Cereb Blood Flow Metab* **44**, 1343-1351 (2024).

16.	Hong, H., Tozer, D.J. & Markus, H.S. Relationship of Perivascular Space Markers With Incident Dementia in Cerebral Small Vessel Disease. *Stroke* **55**, 1032-1040 (2024).

17.	Tournier, J.D.*, et al.* MRtrix3: A fast, flexible and open software framework for medical image processing and visualisation. *Neuroimage* **202**, 116137 (2019).

18.	Tatekawa, H.*, et al.* Improved reproducibility of diffusion tensor image analysis along the perivascular space (DTI-ALPS) index: an analysis of reorientation technique of the OASIS-3 dataset. *Jpn J Radiol* **41**, 393-400 (2023).

19.	Cordero-Grande, L., Christiaens, D., Hutter, J., Price, A.N. & Hajnal, J.V. Complex diffusion-weighted image estimation via matrix recovery under general noise models. *Neuroimage* **200**, 391-404 (2019).

20.	Kellner, E., Dhital, B., Kiselev, V.G. & Reisert, M. Gibbs-ringing artifact removal based on local subvoxel-shifts. *Magn Reson Med* **76**, 1574-1581 (2016).

21.	Graham, M.S., Drobnjak, I., Jenkinson, M. & Zhang, H. Quantitative assessment of the susceptibility artefact and its interaction with motion in diffusion MRI. *PLoS One* **12**, e0185647 (2017).

22.	Pasternak, O., Sochen, N., Gur, Y., Intrator, N. & Assaf, Y. Free water elimination and mapping from diffusion MRI. *Magnetic Resonance in Medicine: An Official Journal of the International Society for Magnetic Resonance in Medicine* **62**, 717-730 (2009).

23.	Csomós, M.*, et al.* Evaluation of the glymphatic system in relapsing remitting multiple sclerosis by measuring the diffusion along the perivascular space. *Magn Reson Imaging* **117**, 110319 (2025).

24.	Taoka, T.*, et al.* Evaluation of glymphatic system activity with the diffusion MR technique: diffusion tensor image analysis along the perivascular space (DTI-ALPS) in Alzheimer's disease cases. *Jpn J Radiol* **35**, 172-178 (2017).

25.	Zhang, W.*, et al.* Glymphatic clearance function in patients with cerebral small vessel disease. *Neuroimage* **238**, 118257 (2021).

26.	Si, X.*, et al.* Neuroimaging evidence of glymphatic system dysfunction in possible REM sleep behavior disorder and Parkinson's disease. *NPJ Parkinsons Dis* **8**, 54 (2022).

27.	Hsiao, W.C.*, et al.* Association of Cognition and Brain Reserve in Aging and Glymphatic Function Using Diffusion Tensor Image-along the Perivascular Space (DTI-ALPS). *Neuroscience* **524**, 11-20 (2023).

28.	Chang, H.I.*, et al.* Gray matter reserve determines glymphatic system function in young-onset Alzheimer's disease: Evidenced by DTI-ALPS and compared with age-matched controls. *Psychiatry Clin Neurosci* **77**, 401-409 (2023).

29.	Zhang, J.*, et al.* Linking white matter hyperintensities to regional cortical thinning, amyloid deposition, and synaptic density loss in Alzheimer's disease. *Alzheimers Dement* **20**, 3931-3942 (2024).

30.	Zhong, J.*, et al.* Unlocking the enigma: unraveling multiple cognitive dysfunction linked to glymphatic impairment in early Alzheimer's disease. *Front Neurosci* **17**, 1222857 (2023).

31.	Han, F.*, et al.* Decoupling of Global Brain Activity and Cerebrospinal Fluid Flow in Parkinson's Disease Cognitive Decline. *Mov Disord* **36**, 2066-2076 (2021).

32.	Yang, H.S.*, et al.* Coupling between cerebrovascular oscillations and CSF flow fluctuations during wakefulness: An fMRI study. *J Cereb Blood Flow Metab* **42**, 1091-1103 (2022).

33.	Fultz, N.E.*, et al.* Coupled electrophysiological, hemodynamic, and cerebrospinal fluid oscillations in human sleep. *Science* **366**, 628-631 (2019).

34.	Jiang, D.*, et al.* Regional Glymphatic Abnormality in Behavioral Variant Frontotemporal Dementia. *Ann Neurol* **94**, 442-456 (2023).

35.	Wang, Z.*, et al.* Reduced coupling of global brain function and cerebrospinal fluid dynamics in Parkinson's disease. *J Cereb Blood Flow Metab* **43**, 1328-1339 (2023).

36.	Raitamaa, L.*, et al.* Spectral analysis of physiological brain pulsations affecting the BOLD signal. *Hum Brain Mapp* **42**, 4298-4313 (2021).
